# The experience of family carers for people with moderate to advanced dementia within a domestic home setting: a systematically constructed narrative synthesis

**DOI:** 10.1101/2023.07.31.23293402

**Authors:** Charles James, Catherine Walshe, Caroline Swarbrick

## Abstract

**Background:** Caring for someone with moderate to advanced dementia within a domestic home setting can be burdensome and time-consuming. To ensure the effectiveness of care planning and delivery, especially towards the end of life, understanding the nature and impact of such caregiving on the family carer is important. Synthesising existing research will allow greater insight into this experience.

**Review question:** ‘How do family carers describe their experience of providing home-based care for people with moderate to advanced dementia?’

**Design:** A narrative synthesis of qualitative research exploring the experiences of people with moderate to advanced dementia and their family carers was conducted. Databases (MEDLINE, CINAHL, EMBASE, PsychINFO, Web of Science and Academic Search Complete) were searched from 1984 to 2020. Similarities and differences between papers were grouped using textual narrative synthesis.

**Findings:** This paper reports findings from a PhD thesis (James, 2021). Included papers (n=17) incorporated those focused on caring for people with advanced dementia (n=8), and those with moderate dementia (n=9). Family carers reported an experience of loss, burden, and grief. Experiences of internal conflict also led to psychological distress. These experiences had a negative impact on the general health of the family carer. Improvement in the family carer’s inter-relationships and the feeling of being useful or having a sense of meaning were reported as positive aspects of caregiving.

**Conclusion:** A complete description of family carers’ experiences may be inadequate in conveying an acknowledgement of dementia as an illness within the domestic home. Their justifications and determinants for balancing family carers’ challenges and distress to morals also remain unclear. Further research is required to ascertain how family carers may proportionally balance their personified loss with their personified value earlier in the disease trajectory.

## Introduction

Accounts of caring for someone living with dementia are mostly rooted within negative experiences, such as the associated burden (Lindeza et al., 2020; Bieber et al., 2019; Farina et al., 2017). Burden, in this respect, may be described as the impact of the difficulties associated with the dementia diagnosis, which increases the level of uncertainty about the future for both the person with the disease and their family (Galvin & Sadowski, 2012; Phillips, 2011). However, not all family carers, who are mostly unpaid, perceive their experiences as burdensome.

Being diagnosed with dementia is often regarded as a burden which increases the level of uncertainty about the future for both the person with the disease and their family (Galvin & Sadowski, 2012; Phillips, 2011). As a factor which also increases the social implications for the people diagnosed and their families, given their difficulty in preparedness, receiving a diagnosis of dementia is often difficult and sometimes described in terms of a feeling of shame and loss of oneself (Xanthopoulou & McCabe, 2019; van Gennip et al., 2016; Lee & Weston, 2011; Aminzadeh et al., 2007).

Life expectancy in dementia varies between individuals. Although a duration of less than 10 years after diagnosis is regarded as a common assumption (Joling et al., 2020), it is unpredictable in comparison with other terminal diseases, such as cancer (Vestergaard et al., 2020; Hall & Sikes, 2018; Arcand, 2015; Harris, 2007). The duration of caring for someone with dementia is therefore likely to last longer than in most other terminal illnesses. The level of decline in dementia is often described in stages, although the duration of each stage is usually unknown. While a cure or effective prevention for dementia is currently unavailable, its stages represent the degree of its severity and are usually identified between the early to the most severe level of decline (Alzheimer’s Society, 2015; Schmidt, 2014; Auer & Reisberg, 1997). A progressive decline in self-care activities of daily living, such as dressing, bathing, and toileting by the person diagnosed is often useful for its stage categorisation (Kumar et al., 2020; Mlinac & Feng, 2016; Marshall et al., 2012).

The responsibilities of family carers usually increase in line with its severity and vary according to the needs of the person cared for (Mesterton et al., 2010). Fulfilling a home home-based care wish in dementia often relies on the willingness of a family carer to take on the role. In some cases, such care provision may be viewed as the family carer’s choice. However, the assumption of the role is mostly associated with a lack of choice (Pertl et al., 2019; Reinhard et al., 2012; Schulz et al., 2012; Al-janabi et al., 2018). A view of family caregiving as performing a moral obligation is therefore plausible.

Given evidence suggests that around 36% of family carers spend more than 100 hours per week, while about 40% provide round-the-clock care (Dementia Carers Count, 2019). In some cases, instances of further unpredictable difficulties, such as behavioural changes are likely over the disease trajectory (van Wijngaarden et al., 2018; Schulz & Eden, 2016). The likelihood of this long caregiving combined with a possible lack of choice leading to family carers’ increased burden and depression is high (Greenwood & Smith, 2019; Karg et al., 2018; Schulz et al., 2012).

Experiences of decline in dementia are unique within its different stages. Within the moderate stage, for example, variations of decline in individual capabilities are possible (Clemmensen et al., 2016; Alzheimer’s Society, 2015). This may be experienced earlier or later by some people, or not at all by others. It is also possible for stages to overlap (Alzheimer’s Society, 2015), which suggests the likelihood of a variation of family carers’ experiences while caregiving within dementia stages. The focus of this review is to explore and synthesise the experiences of family carers while caregiving for people living with dementia within a home setting.

## Aim & objectives

To understand the experiences of family carers who provide care for people with moderate to advanced dementia within a domestic home setting.

## Methods

### Design

A textual narrative synthesis approach was adopted due to its appropriateness in ensuring that similarities and differences between multiple studies were systematically explored (Popay et al., 2006). The stages suggested by Popay et al. (2006), were followed, including identifying the review focus, specifying the review question, identifying studies to include data extraction and quality appraisal, synthesising of the findings, and reporting of findings. The study protocol is published (James et al., 2020).

### Specifying the review question and inclusion criteria

Family carers are people who may not reside with the care recipient, but provide unpaid and significant care (Lindeza et al., 2020; Evans et al., 2019; Woodman et al., 2016). Whilst they are often crucial to the quality of life of the care recipient, they are also regarded as the invisible second patient due to their own care needs (Karg et al, 2018; Sanders, 2016; Brodaty & Donkin, 2009). A purposive search of the key terms, dementia and family carers, and their variations was conducted for the comprehensiveness of the inclusion criteria (Table 1).

**Table 1:**
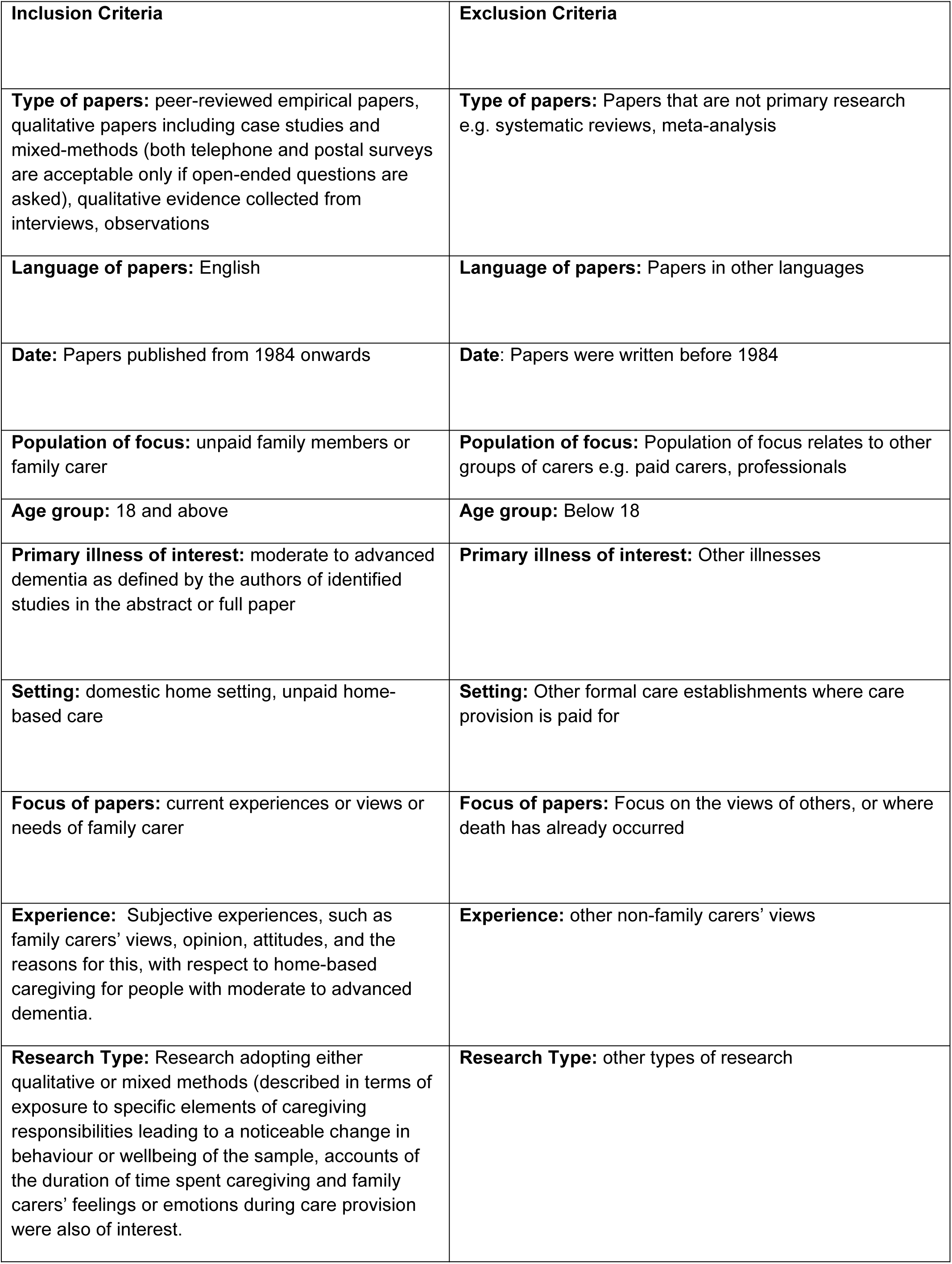
Review inclusion and exclusion criteria.

### Search strategy and process

Searches were conducted in MEDLINE Complete, CINAHL, EMBASE, PsycINFO, Web of Science, and Academic Search Complete. Websites and grey literature were also searched. For all searches, the terms and strategy used a broad range of terms and relevant keywords related to dementia, caregivers, and qualitative studies. These were checked by CW and CS, who were familiar with the domain of dementia and care provision. The search strategy was also tailored for use with each database, using Boolean operators, truncations, and Medical Subject Heading (MESH) as appropriate for each database (Table 2).

**Table 2:**
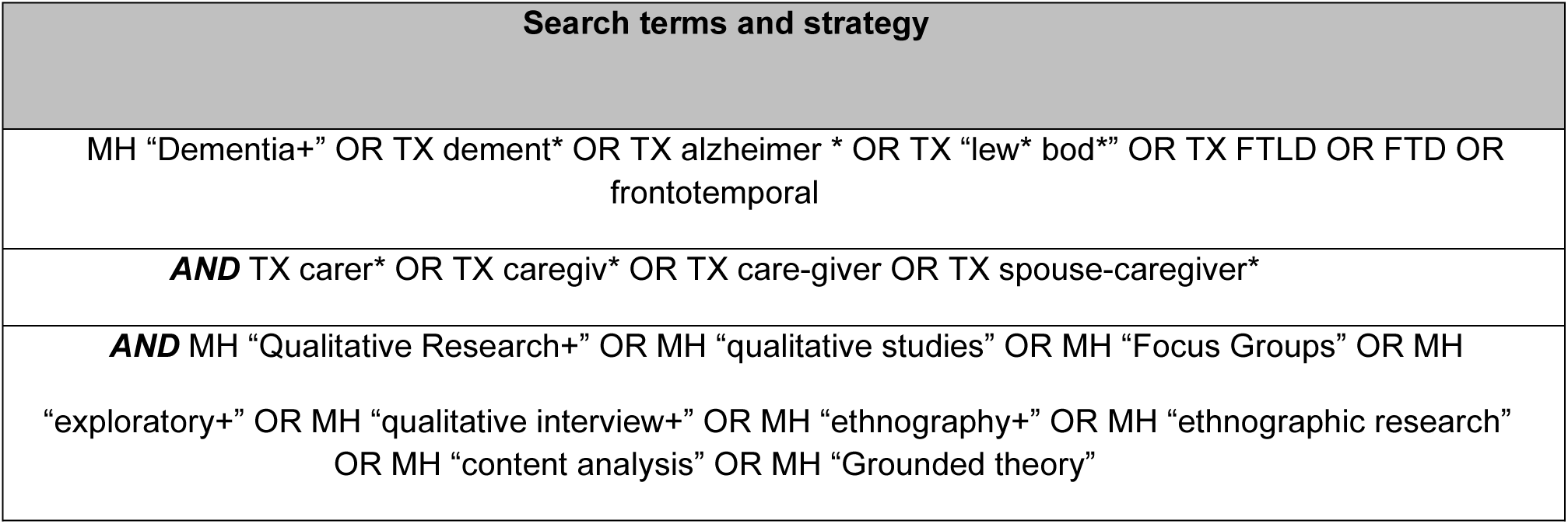
Search terms and strategy.

| Search terms and strategy |
| --- |
| MH "Dementia+" OR TX dement* OR TX alzheimer * OR TX "lew* bod*" OR TX FTLD OR FTD OR frontotemporal |
| **AND** TX carer* OR TX caregiv* OR TX care-giver OR TX spouse-caregiver* |
| **AND** MH "Qualitative Research+" OR MH "qualitative studies" OR MH "Focus Groups" OR MH "exploratory+" OR MH "qualitative interview+" OR MH "ethnography+" OR MH "ethnographic research" OR MH "content analysis" OR MH "Grounded theory" |

The decision to include all papers published from 1984 to 2020 was based on the reported increase in diagnosis and people caring informally (Lewis et al., 2014). Supplementary searches of the reference lists of identified studies were also conducted. The list of studies returned from the searches was imported into Endnote, an online reference management software (Lorenzetti & Ghali, 2013), to remove duplicates and ensure the validity and reliability of the review process’ (Kwon et al., 2015) (See Table 2 above).

### Data extraction

Key data such as author’s name, publication origin and year, setting, population and sample size, aims and objectives, data collection method, and main findings, which show the family carers’ actions were extracted from the included papers.

### Quality and bias assessment

A non-discriminatory tool developed by Hawker et al. (2002) and adapted by Lorenc et al. (2014) was used in assessing the quality of the included papers. Based on the adaptability of its structure to a variety of methodologically distinctive designs, the purpose of this tool was to assess the understanding of quality within the synthesis, rather than to exclude papers of lower quality. The studies included in this review were assessed by two study supervisors, CW and CS. The purpose was to check the results of the appraisal process and ensure the appropriateness of the selected tool as well as the overall process of quality assessment (See Supplementary Table 1).

### Approach to synthesis

A textual narrative synthesis approach was adopted in accordance with guidance from Popay et al. (2006), ensuring the exploration of synthesised words or texts from systematically selected studies using their similarities and differences. A total of 2248 full-text articles were reviewed, with seventeen papers included in the narrative synthesis (Fig 1). Eight studies presented data from those only caring for people with advanced dementia, nine included data from those caring for people with both moderate dementia and advanced dementia. While the review was not focused on establishing a comparison of experiences between genders, it is noteworthy that two studies were dedicated solely to accounts from female family carers (De Silva & Curzio, 2009; de La Cuesta-Benjumea, 2011).

**Fig 1:**
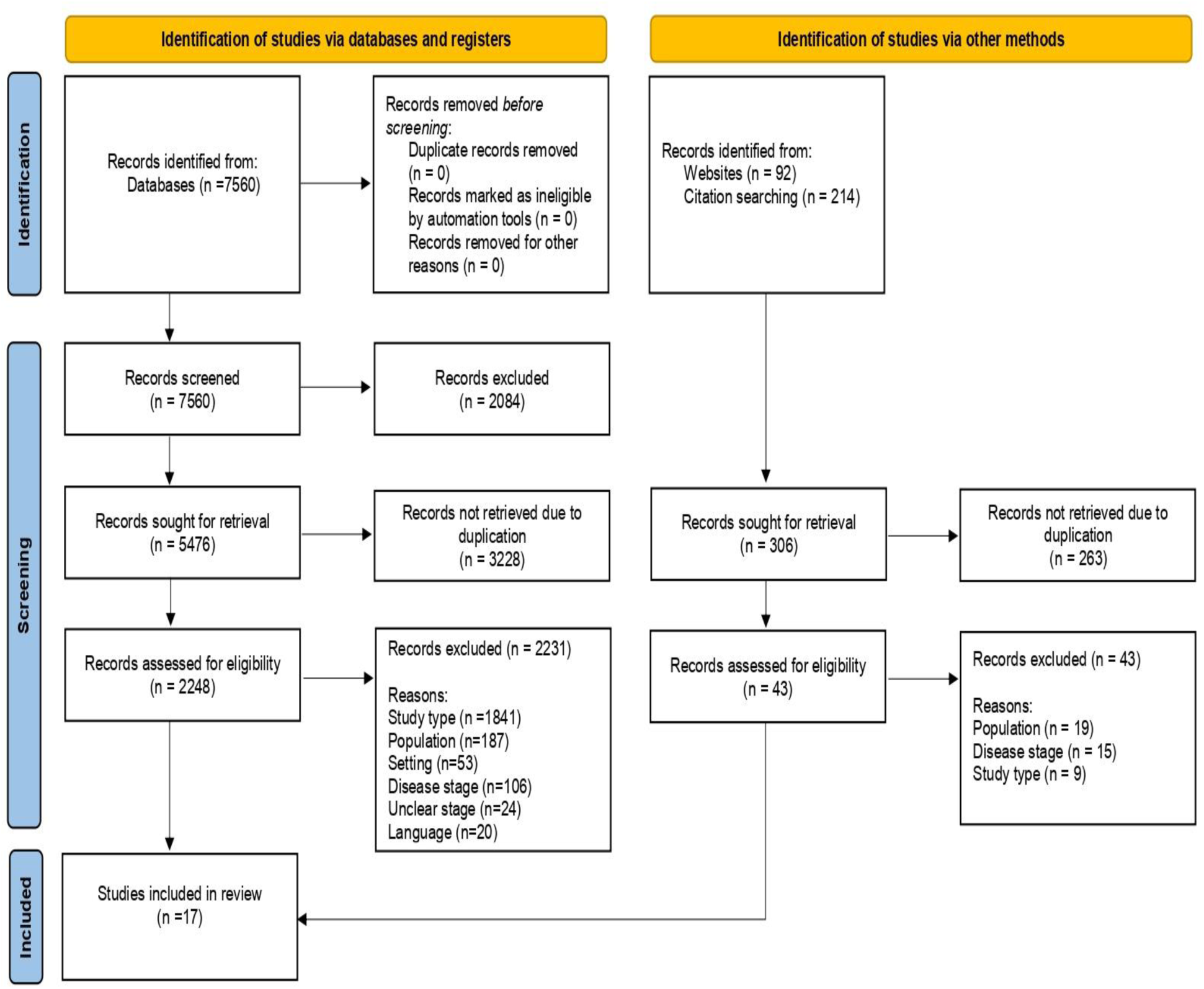
Sources of studies included (PRISMA diagram adapted from Page et al., 2020).

## Review findings

The review findings are presented in three stages. Systematically extracted data are first tabulated to display the similarities and differences between the findings of each selected paper (Popay et al., 2006) (Table 3).

**Table 3:**
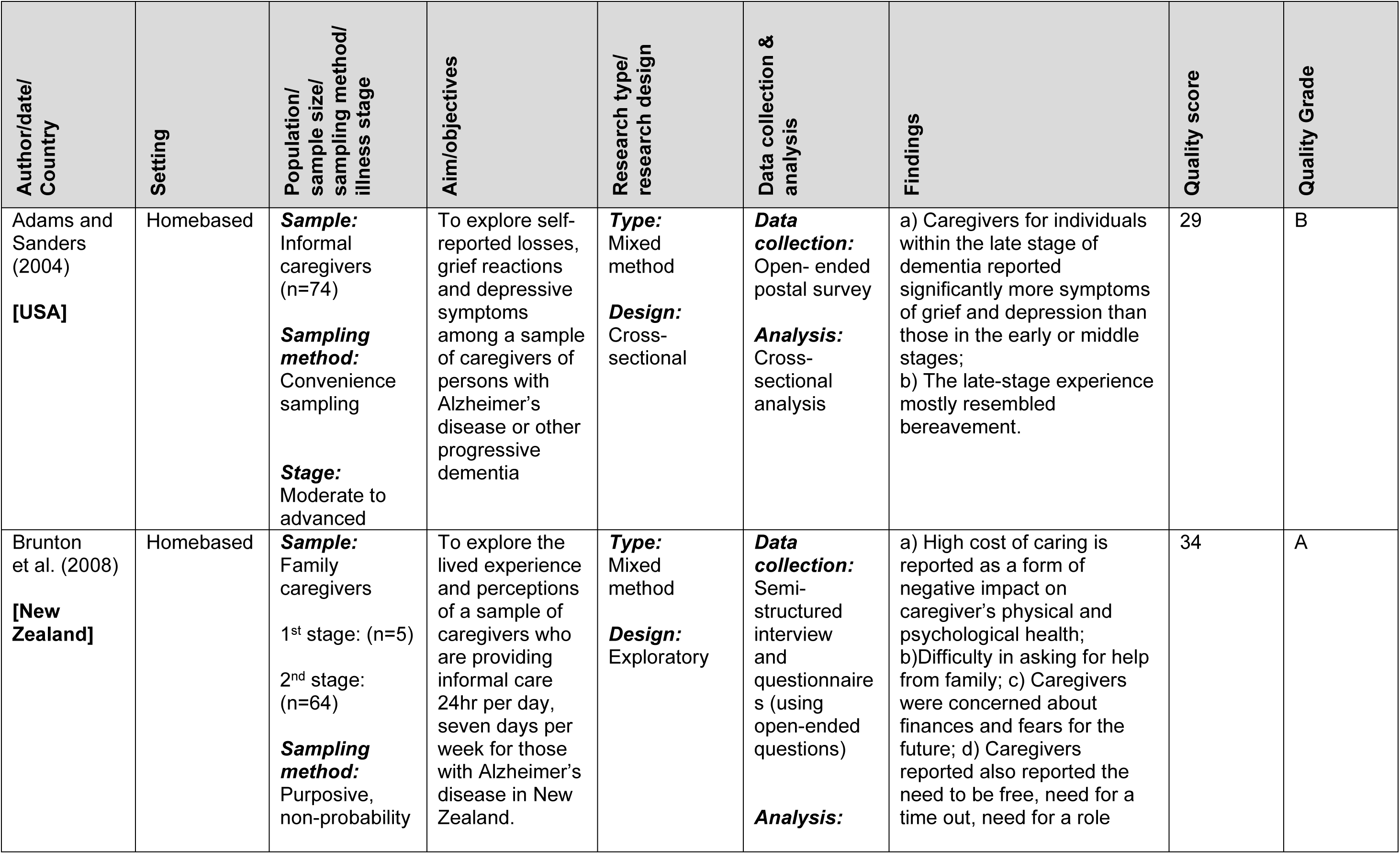
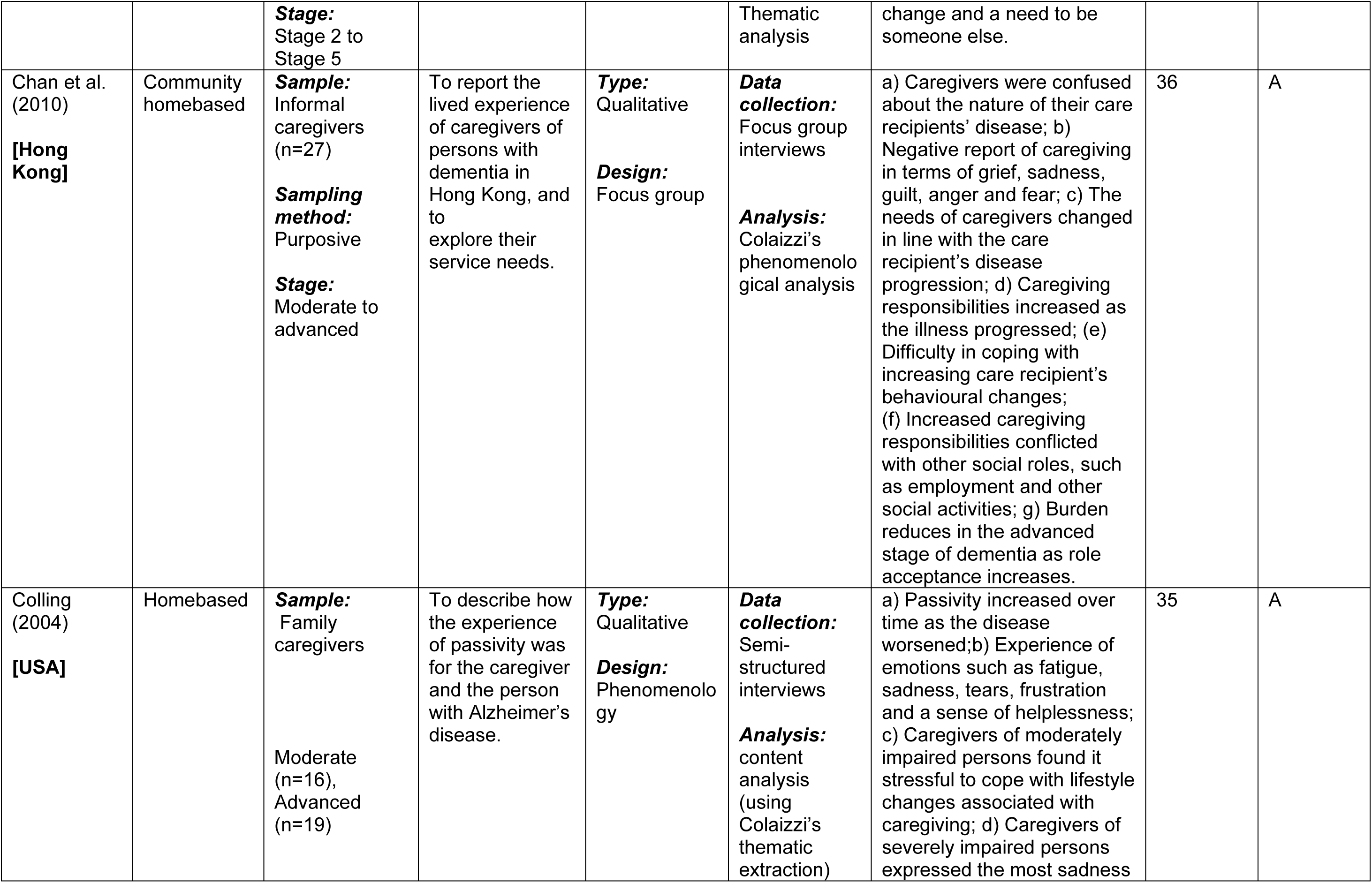
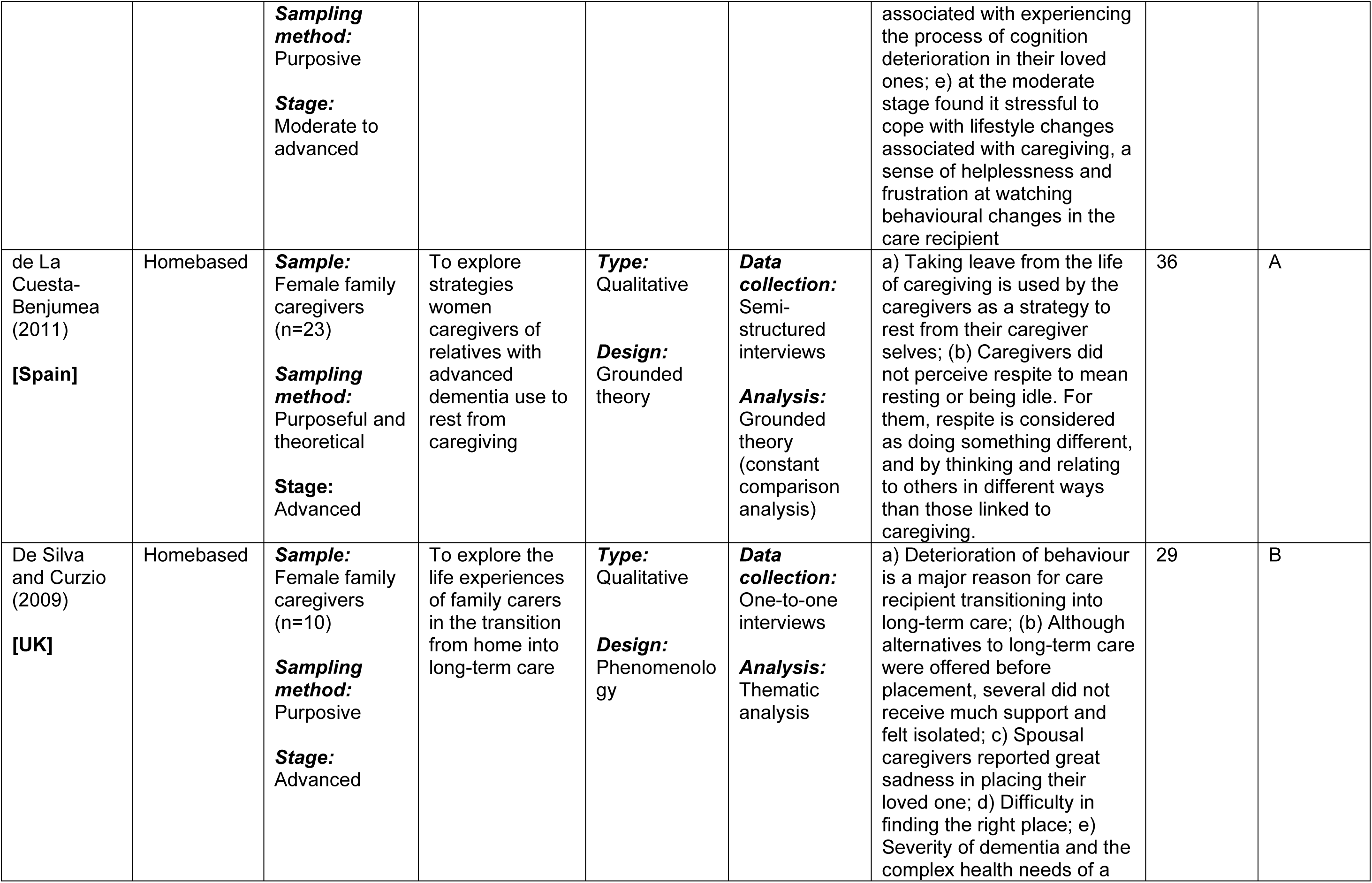
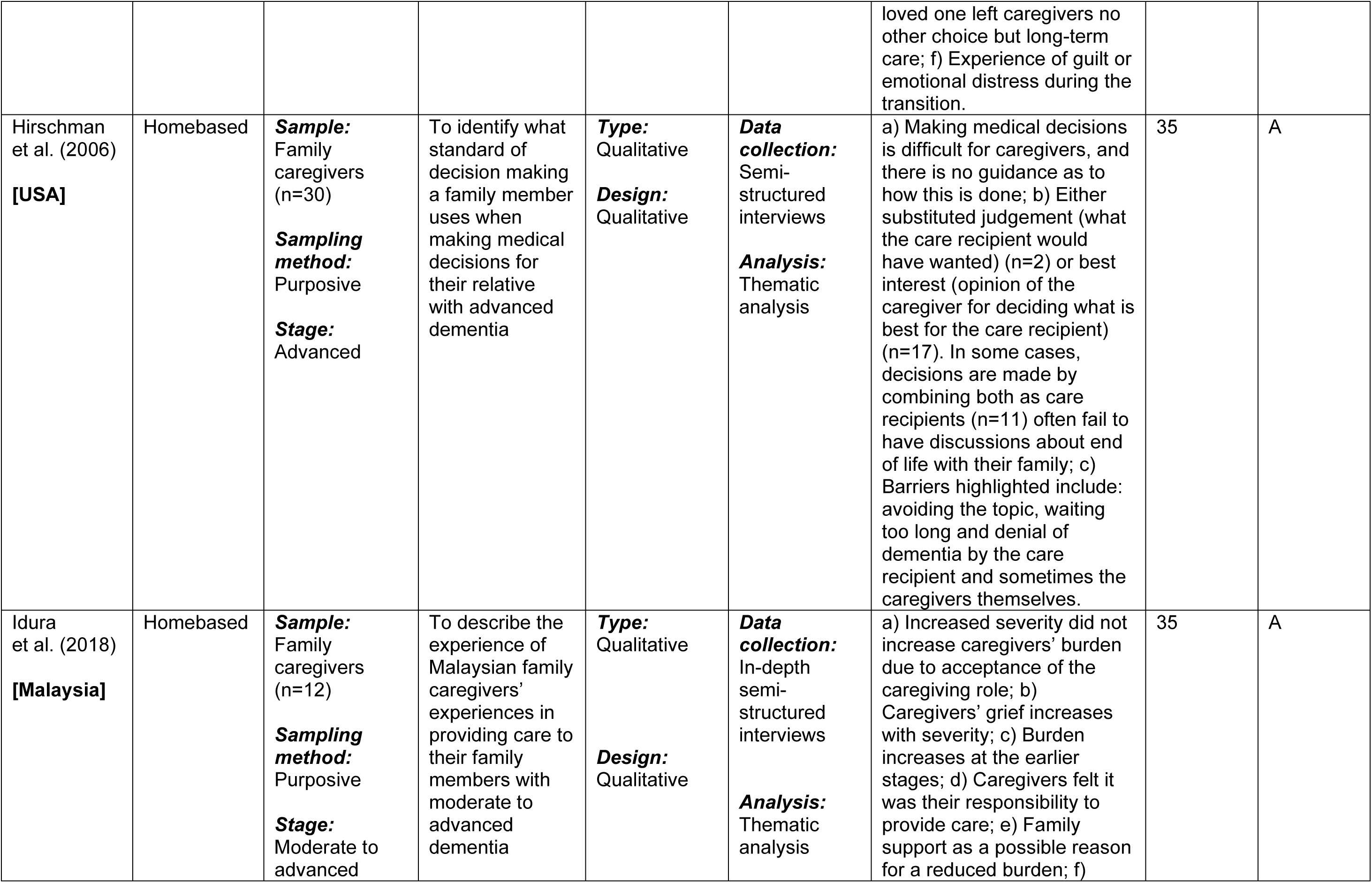
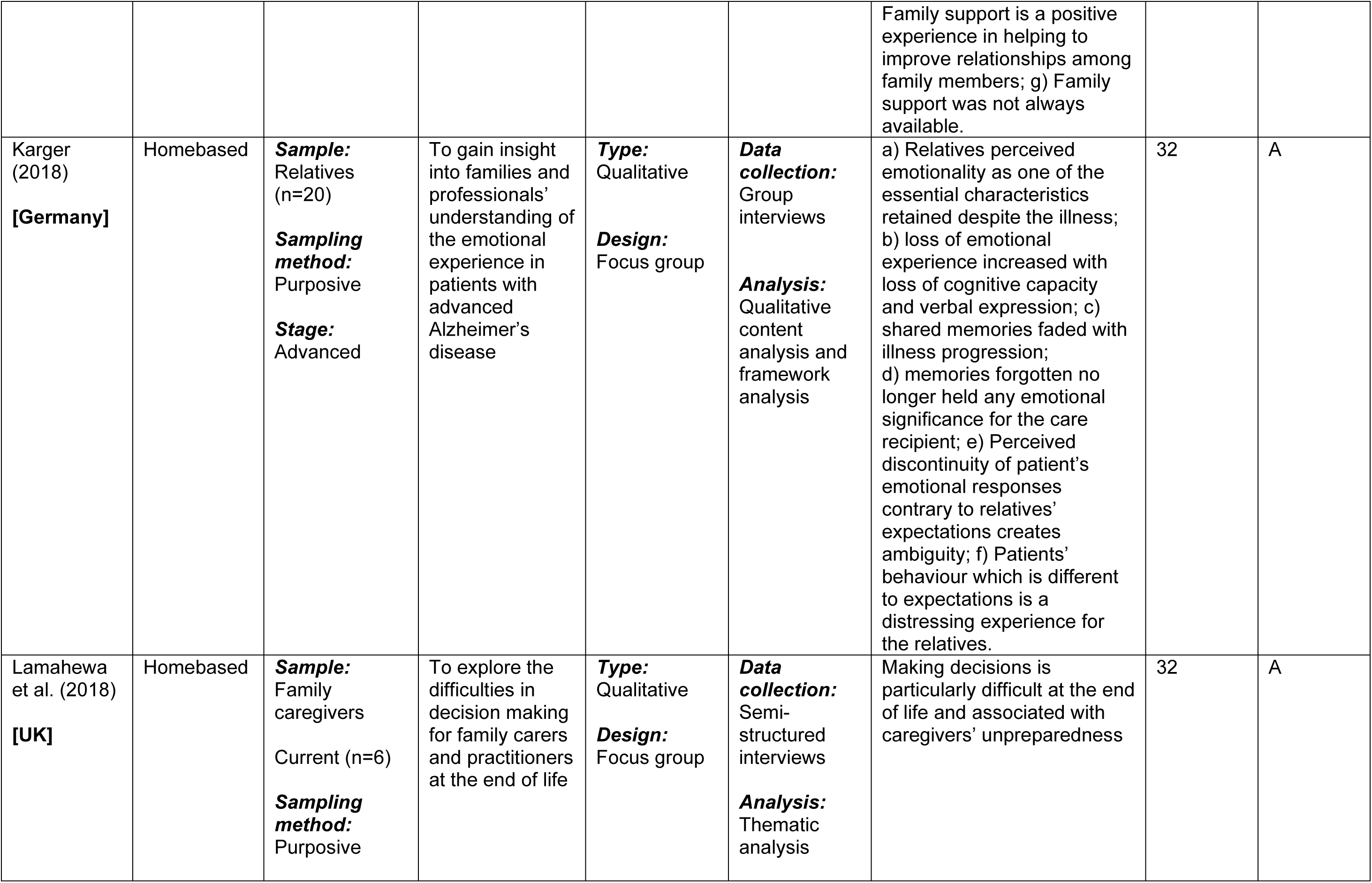
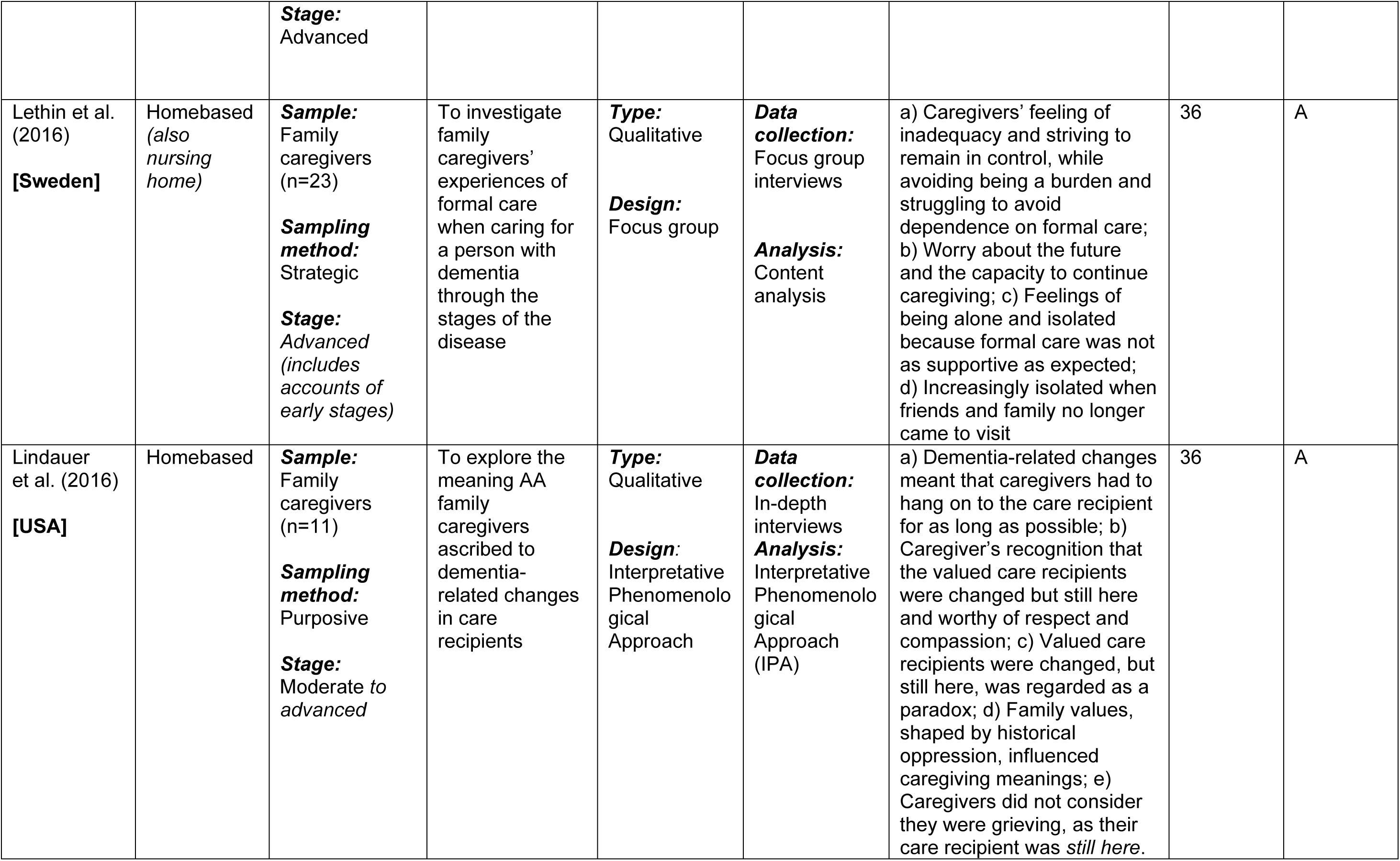
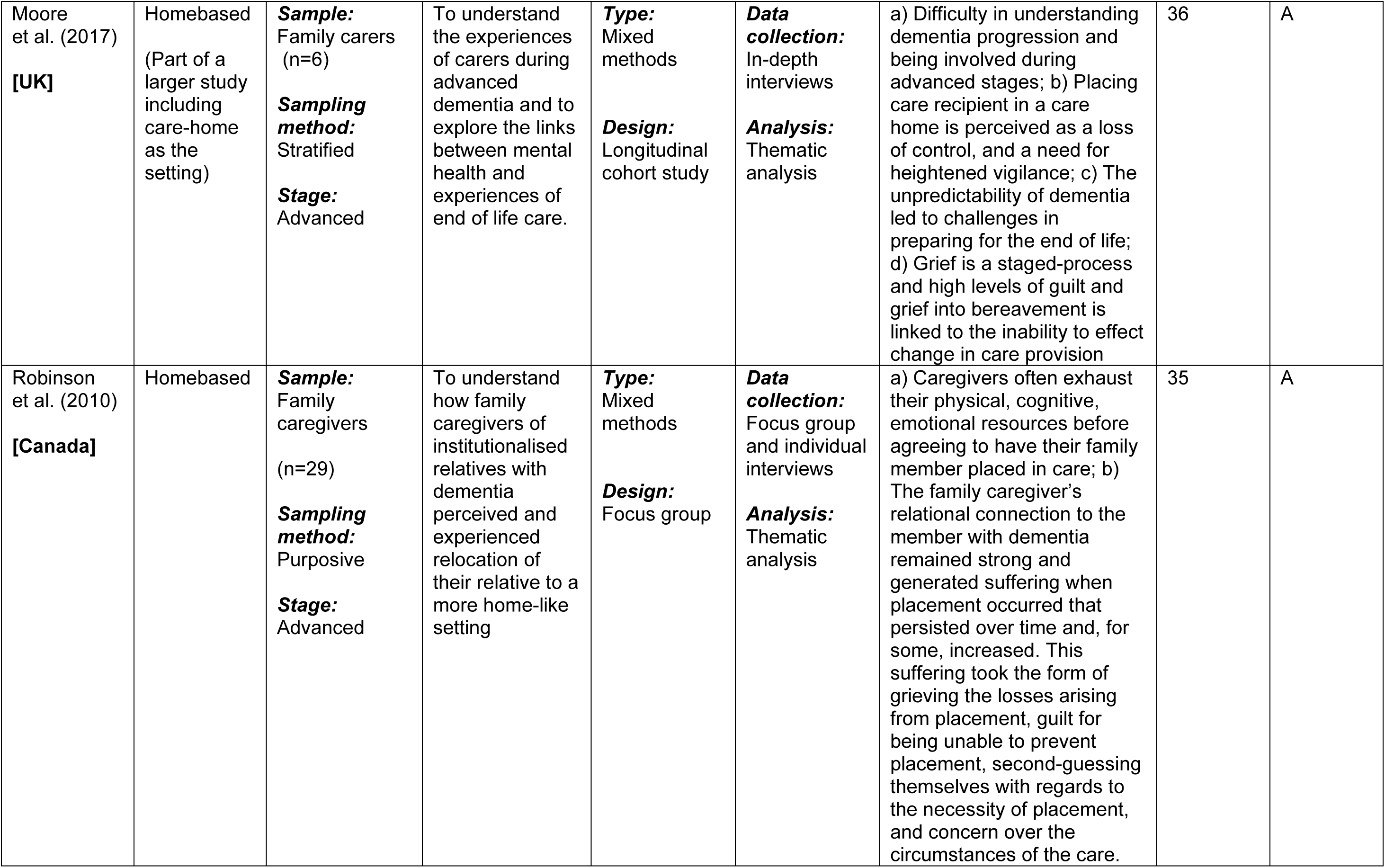
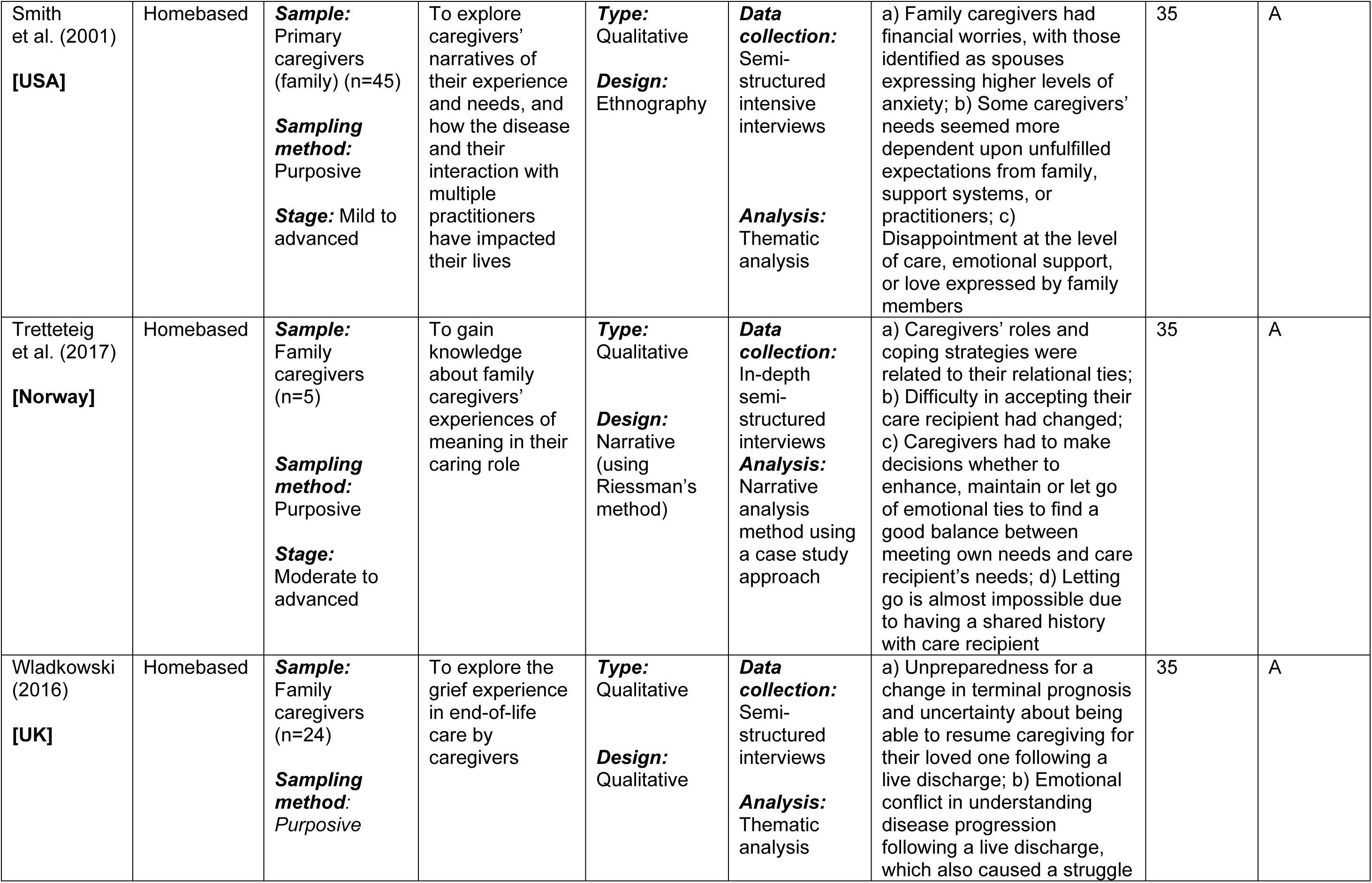
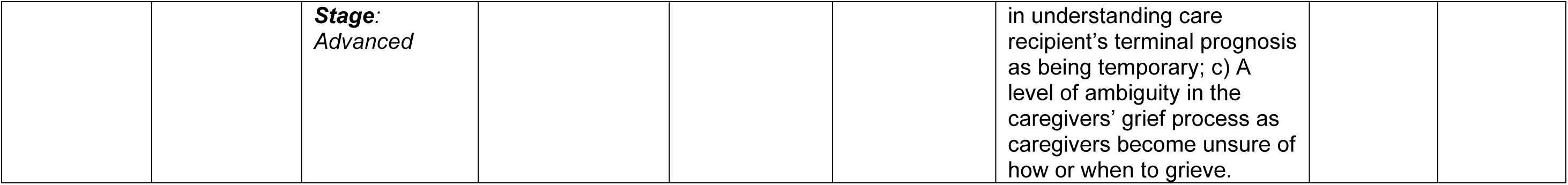
Characteristics of included studies.

## Synthesising the findings

Papers were grouped in the second stage according to the disease stage, country, authors, and population, and the common focus identified in the literature. A textual narrative synthesis of the common focus between the included studies was presented, which shows the interrelationship between the different studies in terms of their commonality on how caregiving experiences differ at both the moderate and advanced stages of dementia (See Supplementary Table 2). In the third stage, a preliminary synthesis is also presented, as a test for the robustness of the synthesised results and as an initial stage to result presentation. Popay et al. (2006) consider this a crucial interrogation of the initial synthesis required to understand the reasons for the findings presented by each study included (See Supplementary Table 3).

## Textual narrative synthesis and presentation of findings

In congruence with Popay et al. (2006), the navigation of the textual narrative synthesis is sequential, showing the family carers’ differentiated accounts of reported burden, loss, grief, how they coped with challenges and the motivation for carrying on with care provision within the explored stages of dementia. These findings are not presented in any hierarchy of importance. However, given its commonality of experience in dementia caregiving, the account of burden is presented first.

### Burden: experience at moderate and advanced stages

Family carers in the moderate stage experienced increased caregiving responsibility due to increased care recipient’s needs (Idura et al., 2018; Chan et al., 2010; Adams & Sanders, 2004). Extended time was spent caregiving as a result, which led to a clash with other social roles and a difficulty in meeting the family carers’ own needs (Idura et al., 2018; Chan et al., 2010). In the case of spousal caregivers, extended time spent on caregiving was also highlighted (Idura et al., 2018; Chan et al., 2010; Adams & Sanders, 2004). Whilst the extended time caregiving was considered a necessity in some cases as highlighted by Adams and Sanders (2004), the reason for this was not given. Swall et al. (2019) and Hellström et al. (2005) suggested, however, that this may be attributable to their couplehood.

Increased caregiving responsibility caused family carers’ increased burden at the moderate stage of dementia (Adams & Sanders, 2004). Their difficulty in meeting own needs also intensified over time (Chan et al., 2010). This finding represented a high cost of caring with possible risk to family carers’ mental and physical health (Idura et al., 2018; Brunton et al., 2008; Adams & Sanders, 2004). Furthermore, contrary to the report of an increase in caregiving being the only association to caregivers’ increased burden, evidence also showed that family carers’ increased burden was linked to progressive decline and changes in the care recipient’s behaviour (Idura et al., 2018; Karger, 2018; Lindauer et al., 2016; Chan et al., 2010).

The collective account at the advanced stage further associated family carers’ burden with care recipient’s behavioural changes, which existed as part of progressive decline in dementia (Colling, 2004). Passivity, a distinctive change in care recipient’s behaviour, which also affects functionality in thinking, movement, and interactions with others, was cited as an example (Colling, 2004). However, the severity of this change did not increase the family carers’ burden (Idura et al., 2018; Chan et al., 2010). Passivity also changed the care recipient’s emotional responses and caused the family carers’ difficulty in accepting care recipients’ changes (Karger, 2018). Overall, as the care recipient’s passivity increased, the family carers’ helplessness, sadness, and frustration at the inability to help also increased (Colling, 2004).

Passivity was, however, not only distinctive at the advanced stage. Common symptoms of increased agitation, aggression, and withdrawal were also reported at the moderate stage (Lindauer et al., 2016; Chan et al., 2010; Colling, 2004). As care recipients became more passive and required help with functional activities such as personal care, the demand for their carer’s attention also increased (Colling, 2004). This also caused the family carers’ burden to increase in line with care recipients’ increasing passivity at this stage of the disease.

Some family carers found it difficult to accept the process of care recipients’ decline and expressed the most sadness at the advanced stage (Colling, 2004). Despite this, they accepted their role better, as they were adjusted to their new lifestyle (Chan et al., 2010; Colling, 2004). In comparison, less burden was reported by family carers at the moderate stage (Colling, 2004), which was attributed to their reliance on external resources such as extended family support (Idura et al., 2018; Chan et al., 2010; Brunton et al., 2008). While the availability of external support was commonly perceived as being able to reduce burden by family carers for recipients at both stages of dementia (Idura et al., 2018; Chan et al., 2010), evidence of consistency in burden reduction at either stage was not discussed in the reviewed literature.

Some family carers were assisted by their extended family (Brunton et al., 2008; Smith et al., 2001). Others experienced difficulty in asking for support (Chan et al., 2010; Brunton et al., 2008; Smith et al., 2001). Some family carers had to *“apply pressure”* to receive any assistance at all (Brunton et al., 2008), and disappointment was sometimes experienced at the level of assistance offered (Smith et al., 2001). Although some positive experiences were reported, such as the report of an improvement in interrelationships (Idura et al., 2018; Chan et al., 2010), tension within the family was a possibility (Chan et al., 2010). This also supports the finding that the reduction of family carer’s burden through extended family support at both stages of dementia was not true in all cases.

Poor understanding of family carers’ needs (Idura et al., 2018; Chan et al., 2010), was commonly evidenced as one of the factors for inadequate familial assistance at both stages (Chan et al., 2010; Brunton et al., 2008). Other reasons include living away from the care recipient (Chan et al., 2010; Brunton et al., 2008), and familial indifference (Chan et al., 2010; Brunton et al., 2008). Receiving assistance from the family was therefore not always available. Moreover, it was possible for family expectations to attach a sense of duty to caregiving (Idura et al., 2018; Tretteteig et al., 2017; Chan et al., 2010). As such, caregiving therefore became an obligation at both stages (Idura et al., 2018; Chan et al., 2010).

### Loss: experience at moderate and advanced stages

Family carers’ loss was explored from two perspectives. The first, mostly descriptive of the narrative at the moderate stage, was related to their personal changes. This was described in relation to own time, freedom, independence, and being able to socialise (Idura et al., 2018; Chan et al., 2010; Brunton et al., 2008; Adams & Sanders, 2004). These lifestyle changes were stressful (Colling et al., 2004), and attributable to their increased caregiving responsibilities (Chan et al., 2010; Adams & Sanders, 2004; Colling, 2004). It was further reported by those who identified themselves as spouses, that this type of loss was the highest, especially in cases where caregiving ceases to be provided at home at the advanced stage of the disease (Adams & Sanders, 2004). Their loss was also accompanied by guilt (Adams & Sanders, 2004).

The second perspective was associated with the care recipient’s changes, mostly at the advanced stage. This was described as the care recipient’s altered behaviour (Colling, 2004), and the difference over time (Adams & Sanders, 2004; Colling, 2004). For this type of loss, care recipients were described in relation to who they once were (Lindauer et al., 2016; Adams & Sanders, 2004), in terms of their personality (Tretteteig et al., 2017; Lindauer et al., 2016; Adams & Sanders, 2004), and relationship once jointly shared with the family carer (Tretteteig et al., 2017; Lindauer et al., 2016; Brunton et al., 2008; Adams & Sanders, 2004).

Some family carers did not emphasise their care recipient’s personality changes (Lindauer et al.,2016). For them, the care recipient’s significance or value was instead considered of more relevance (Lindauer et al., 2016), than the changes attributed to what has been lost to dementia (Lindauer et al., 2016). These family carers therefore held on to *“what remained”* of the care recipient for as long as possible (Lindauer et al., 2016, p.738). However, some family carers placed more emphasis on their care recipient’s lost personality (Adams & Sanders, 2004), instead of what remained (Lindauer et al., 2016). In comparison to those who considered the value of their care recipients to be of more significance, these carers regarded their care recipients as already gone (Adams & Sanders, 2004).

A commonality in the description of loss was therefore established as the recognition of the absence of the care recipient’s valued personality (Lindauer et al., 2016; Adams & Sanders, 2004). Both of these studies also gave evidence of a common experience of a paradoxical feeling, described as holding on to ‘what remains of the care recipient’ (Lindauer et al., 2016), or holding on to ‘someone who was already gone’ (Lindauer et al., 2016; Adams & Sanders, 2004). These two expressions appear synonymous. However, while evidence of historical hardship was cited between carers and care recipients in Lindauer et al. (2016), the existence of such a relationship was not described by Adams and Sanders (2004).

### Grief: experience at moderate and advanced stages

Across the trajectory of dementia, variance in grief level existed (Lindauer et al., 2016; Adams & Sanders, 2004). As an emotional response to losing a loved one (Moore et al., 2017; Adams & Sanders, 2004; Colling, 2004), its severity was described as being at its highest at the advanced stage (Idura et al., 2018; Wladkowski, 2016; Adams & Sanders, 2004). In some cases, while family carers’ perception of grief was regarded as part of the pre-death experience (Moore et al., 2017; Lindauer et al., 2016; Adams & Sanders, 2004), it was also associated with the observation of the care recipient’s decline (Colling, 2004). Irrespective of the relationship between the family carer and their care recipient, the association between higher grief level and higher loss reported at the advanced stage was unclear.

Some family carers reported a no-grief experience at both stages of dementia. At the moderate stage, for instance, this was considered a response to the delay in the care recipient’s decline (Adams & Sanders, 2004). As a result, carers at this stage concentrated on their care recipient’s lost personhood as an outlet for their perceived loss (Adams & Sanders, 2004). Although this claim was also supported by Lindauer et al. (2016), both studies differed in some of their findings. While Lindauer et al. (2016) asserted that carers who reported a no-grief experience focused on their care recipient’s personality which remained, Adams and Sanders (2004) reported that family carers’ emphasis was on their care recipient’s already lost personality. The carers’ no-grief experience was therefore related to anger, self-pity, or shame. (Adams & Sanders, 2004). On the other hand, the no-grief experience reported, especially by those with a shared history of hardship (Lindauer et al., 2016), establishes a defining characteristic of a shared identity.

Holding on to what remains caused some family carers’ unpreparedness for the care recipient’s death (Moore et al., 2017; Lindauer et al., 2016), as caregiver-attributed value was preserved longer. Indeed, for some, grief was irrelevant, and the reality of actual death was unacceptable (Lindauer et al., 2016). In this situation, a prolonged after-death grieving was therefore possible.

### Dealing with challenges and continuing with caregiving

Caregiving was perceived as a natural obligation by some family carers (Lindauer et al., 2016), and they used religion to cope with the caregiving challenges encountered (Idura et al.,2018; Lindauer et al., 2016; Colling, 2004; Smith et al., 2001). Through religion, caregiving was perceived by some as rewarding or as the ‘inheritance of good morals’ (Idura et al., 2018), while others considered it as fulfilling an important role (Lindauer et al., 2016), or doing the right thing (Tretteteig et al., 2017). These perceptions were common across both stages of the disease.

Caregiving expectations carried a sense of obligation, attributed by societal and cultural values (Idura et al., 2018; Chan et al., 2010), or the carer’s self-induced expectations to provide care (Idura et al., 2018; Tretteteig et al., 2017). This, however, varied with relationship between the carer and the care recipient (Idura et al., 2018; Tretteteig et al., 2017; Chan et al., 2010). In some societies, for instance, the obligation to provide care to older people was placed on their family (Lindauer et al., 2016; Chan et al., 2010) by sources such as the society (Tretteteig et al., 2017), religion (Idura et al., 2018), culture (Chan et al., 2010), a sense of history, and identity shared with the care recipient (Lindauer et al., 2016). In these societies, the perceived sense of duty, therefore, influenced caregiving (Tretteteig et al., 2017).

The fear of inadequacy in knowledge and skills existed in some family carers’ preparation for their increased caregiving responsibilities (Idura et al., 2018). At the moderate stage, this was managed by finding a balance between own expectations and the reality of the care recipient’s behavioural changes (Idura et al., 2018). Although some carers used their prior knowledge of the care recipient’s background for managing challenges faced in caregiving (Colling, 2004), others considered care recipient’s placement in a care home (Lethin et al., 2016; Adams & Sanders, 2004; Smith et al., 2001), or receiving support at home (Lethin et al., 2016). For most carers, placing the care recipients away from home was however inconceivable, especially at the advanced stage.

A preference for continuing with home-based caregiving was highlighted, especially in situations where the care recipient’s decline necessitates consideration for care provision outside of the home setting (Lindauer et al., 2016). Some family carers found this decision difficult, and the wish to maintain control on caregiving persisted (De Silva & Curzio, 2009). This wish to maintain control increased their ability to make meaning of the caregiving role (Tretteteig et al., 2017). Although this was considered a characteristic of carer’s grief (Adams & Sanders, 2004), maintaining control was also crucial for balancing the carer-to-recipient relationship.

Loss of control was accompanied by a feeling of guilt and sadness, especially where the care recipient’s placement away from home was necessary (Moore et al., 2017; Tretteteig et al., 2017; De Silva & Curzio, 2009; Adams & Sanders, 2004; Chan et al., 2010). The fear of poor caregiving by others outside of the home setting was cited as a reason (Moore et al., 2017). Situations occurred whereby some of the family carers’ guilt and sadness persisted beyond bereavement, due to self-blame (Moore et al., 2017). Although evidence was given which showed that this further increased the reluctance to seek outside care away from home, the influence of family carers’ perceived obligation to continue caregiving, and their need to maintain control in such cases was unclear.

Moving the person cared for away from home was difficult (Lindauer et al., 2016). Some family carers used respite as their preferable means for managing their caregiving-related challenges (Idura et al., 2016; Lethin et al., 2016). Generally, respite is used in briefly taking away caregiving responsibilities (de La Cuesta-Benjumea, 2011). Variation in its description, however, exists within the reviewed literature. According to Brunton et al. (2008), respite is described as a physical location, where care recipients reside in order for their carers to have a break, or a situation where caregiving responsibility is shifted to another person, such as a family member (Idura et al., 2018; Brunton et al., 2008). However, de La Cuesta-Benjumea (2011) described respite as a situation whereby family carers may separate themselves from their caregiving identity and briefly assume a different identity (de La Cuesta-Benjumea, 2011). This therefore suggests that achieving rest is possible without the need for family carers’ physical separation from their care recipients.

Alternative identities were achievable through mental isolation, whereby caregiving responsibilities were mentally paused by the family carer to give them an opportunity to explore other identities (de La Cuesta-Benjumea, 2011). As such, managing caregiving challenges was possible, regardless of the carer’s physical location (de La Cuesta-Benjumea, 2011). This may be useful in situations where a respite facility is unavailable or where there is a reluctance to place the care recipient away from home.

## Discussion

The central focus of experiences was mainly negative, and broadly on family carers’ accounts of loss, burden, and grief. These experiences are reported in the wider literature as attributes commonly associated with general experiences of dementia caregiving. Some positive aspects of caregiving were also reported, including improvement in family interrelationships, and the family carers’ feeling of being useful and having a sense of meaning. An association between loss, burden, and grief of the caregiver exists, according to Meuser and Marwit (2001). The experience of these was not descriptive of all family carers’ narratives in this review. Furthermore, these are not the only dementia-caregiving related experiences, as a variety of other experiences, such as stress, anxiety, and depression are also reported in the wider literature (Blandin & Pepin, 2017; Chan et al., 2010; Adams & Sanders, 2004). The order of these experiences by the family carers, however, remains unclear.

Narratives of loss were explored from two different perspectives in this review. Firstly, it was described in relation to family carers’ unmet needs, as a loss of social roles and difficulty in fulfilling own needs. This was termed ‘*personified loss’,* as descriptions represented the family carers’ perception of own loss. The description of loss in terms of family carers’ own needs has been given in other studies. Brodaty and Donkin (2009) describe this in terms of social isolation which may result in a loss of meaning in life. Similar findings have also been reported in this current review, and it is suggested that this experience may worsen over time due to the possibility of emotional burden exposure from increased caregiving (Greenwood et al., 2019; Brodaty & Hadzi-Pavlovic, 1990).

Changes to the way of life were described by the family carers’ increased burden through caregiving responsibilities. Inability to socialise at this stage of dementia was viewed as a conflict between increased psychosocial distress and associated changes to caregiving focus. In relevance to this experience, the description of family carers’ *personified loss* may therefore be a precursor to a *personified cost* in terms of opportunities forfeited for accepting caregiving responsibilities. Hence, the likelihood of family carers’ *personified loss* being interpreted as a forfeited way of life rather than expressed as part of the normal progression of dementia caregiving is elevated.

Secondly, loss was also described as a caregiver-attributed value in relation to the care recipient’s changes and termed ‘*personified value’* in this review. Dupuis (2002) identifies this as a detrimental attribute due to the possible closeness between the carer and the person cared for. This type of loss was further described in terms of ambiguity given that it acknowledges the absence of a valued personality (Dupuis, 2002; Boss, 1999; Jones & Martinson, 1992). In similarity to the findings by Dupuis (2002), evidence by Boss (1999), and Jones and Martinson (1992) suggest that family carers experience a state of loss where the person cared for was considered gone while still physically present. Similarity of findings in this review shows that although loss suggests a *personified value*, the view of the person cared for as ‘already gone’ while still alive was also shared.

The possibility of increasing family carers’ emotional hold to the personalities that remain is considerable, given the fear of losing *personified value.* Subsequently, holding on persists longer as a paradox where balancing *“what remains”* of the care recipient with *“what is already lost”* becomes difficult. As in other terminal conditions, evidence suggests that this experience, also referred to as anticipatory grief, is a pre-death response to the process of progressive decline in dementia (Pérez-González et al., 2021; Cheung et al., 2018; Blandin & Pepin, 2017; Meuser & Marwit, 2001). Thus, in cases where the realisation of dementia contributes to the early attribution of *personified value*, family carers’ fear of losing *what remains* may cause a higher level of grief in both stages of dementia.

The resolution of grief corresponds with family carers’ acceptance of own loss (Prigerson & Maciejewski, 2008). As an *“emotional inability to accept the loss of something cherished”,* non-acceptance results in grief and attributes such as guilt, frustration, and helplessness (Prigerson & Maciejewski, 2008, p.435). While grief may be described as a natural response to family carers’ loss, those who report higher levels of pre-death grief have a higher risk of health complications post-death (Shuter et al., 2014; Chan et al., 2010; Givens et al., 2011; Rando, 2000). The risk of health complications from earlier stages of the disease and possibly post-death may therefore increase. An early resolution of caregiver-attributed *personified value* is therefore necessary.

Grief was, however, not an experience reported by family carers who shared a historical hardship with the care recipient in this review. In commonality with other caregivers, holding on to *personified value f*or longer was highlighted. While it is appropriately assumed that “*something cherished”* (Prigerson & Maciejewski, 2008, p.435) may refer to factors such as family carers’ own time, freedom, or care recipient’s value, as well as the identity shared as a result, the loss described may also be personified. Further research is, however, required for differentiating between a caregiver-attributed value of what remains and what is already lost to dementia, and its influence on the possible duration of prolonged after-death grieving.

It is further appropriate to also consider that, for most family carers, a grief-experience is reported through both stages of dementia irrespective of their relationship with the care recipient (Meuser & Marwit, 2001). As the care recipient declines, *personified loss* increases in line with the increased demand for the carers’ attention. Consequently, loss of *personified value* and family carers’ grief may also increase at both stages. The justification and determinants for balancing *personified value* against family carers’ *personified loss* are however, currently unclear. Given these findings, the issue of balancing family carers’ reported experiences against their reasons for continuing caregiving at both stages of the disease therefore arises. It is assumed that other factors may also be present in the family carers’ experiences, whereby powerlessness is felt in how future priorities are perceived and how the balance between *personified value* and *personified loss* may influence continued caregiving. This also calls for an understanding of a possible distress at choosing the right course of action to take in such situations (Oh & Gastmans, 2015; Silén, 2011; Jameton,1993; 1984).

Some gaps in current knowledge were also identified during this review. It was claimed that variation in family carers’ grief-experience across dementia stages increases with severity (Idura et al., 2018; Lindauer et al., 2016; Wladkowski, 2016; Adams & Sanders, 2004).

Moreover, a higher level of grief-experience is associated with the advanced stages (Adams & Sanders, 2004). Clarity is however required to determine whether a higher grief level at the advanced stage is resultant from a higher loss experienced from earlier stages.

Placement of caregiving expectations from different sources was suggested as a reason for an induced sense of obligation on family carers (Idura et al., 2018; Tretteteig et al., 2017; Chan et al., 2010). This varied according to the relationship between the family carers and their care recipients (Idura et al., 2018; Tretteteig et al., 2017; Chan et al., 2010), and serves as a factor which influenced behaviour (Tretteteig et al., 2017). While the possibility of negative outcomes was reported, the relationship between the family carers’ obligation and grief level remained unexplored. Although similarity of experience of holding on to care recipients’ personality was evidenced in the literature, an ambiguity remained between clear definition of what remains of the care recipient (Lindauer et al., 2016) and what was already lost to dementia (Adams & Sanders, 2004), as well as its influence on determining the duration of prolonged after-death grieving.

## Implications

The impact of higher levels of pre-death grief in caregiving for someone with dementia is widely recognised in association with health complications post-death (Shuter et al., 2014; Chan et al., 2010; Givens et al., 2011). Although higher grief level is commonly attributed to the advanced stages, the possibility of attributing a *personified value* earlier in caregiving is evident from this review. For the family carer, the fear of losing care recipients’ *personified value* carries a higher level of grief in both stages, thereby increasing the risk of health complications earlier and possibly post-death. This may influence how home-based caregiving is viewed by family carers and their willingness to accept or continue caregiving responsibilities at home. Given a projected increase to over two million dementia family carers by 2051 (Knapp et al., 2014), the implication is therefore severe for sustaining home-based caregiving for people with moderate to advanced dementia in the UK.

## Strengths and limitations of the review

The adoption of a narrative synthesis approach allowed for heterogeneity of evidence in this review. This approach may have contributed to the robustness of synthesised evidence. A difficulty in managing the scope of possible evidence was however high, as a large number of individual papers were checked for the inclusion criteria to be met. Many papers were found where the population of interest was either missing or incorrectly reported. This difficulty was further exacerbated by the inaccurate classification of some papers by their authors, thereby making both the screening stage and inclusion problematic from reading their abstracts. Variation of accounts contributes to how family carers’ experience of loss may be explored in future research. Given that some accounts used were extrapolated from a wider context of discussion within the reviewed literature, full justification of accounts may therefore be difficult (Polit & Beck, 2010). Generalisation of the findings at either of the moderate and advanced stages of dementia may also be difficult. However, the main purpose of any qualitative endeavour, such as this review, is to provide a rich understanding of people’s accounts. Polit and Beck (2010) highlight that the transferability of these accounts across clearly defined settings and contexts is therefore appropriate.

## Conclusions and recommendations

Family carers’ descriptions of experiences at the moderate to advanced stages of dementia vary. Overall, accounts suggest that spending time with the person cared for enhances the acknowledgement of the illness experience by the family carer. Using a narrative synthesis approach, the findings of this review suggest that the level of understanding of the illness experience differs. A complete description of all experiences may therefore be inadequate in conveying an acknowledgement of the illness within home-based caregiving. Given accounts of burden, loss, grief, and dealing with challenges in making meaning show that increasing caregiving responsibilities increased the family carer’s difficulties, especially at the moderate stage. Understanding the experience of loss through to the advanced stages is also crucial. It is suggested that the caregiver-attributed *personified value* resulted in holding on to what remains of the care recipient for longer. Research is however required to understand whether this may lead to a higher grief experience from the moderate stage or earlier. Further research is also required to explore how family carers may proportionally balance their *personified loss* with their *personified value* earlier in the disease trajectory, given that the justification and determinants for balancing these, and whether distress to morals is felt, remains unclear.

## Supporting information

Supplementary Table 1: Result of the quality assessment of included studies

Supplementary Table 2: Textual narrative synthesis exploring relationships between studies

Supplementary Table 3: Preliminary synthesis of studies

## Acknowledgement

We would like to express our gratitude to Professor Katherine Froggatt for her advice.

## Ethics statement

Ethical approval was not required for this review.

## Data availability

Data supporting this review are included within the paper.

## References

Adams, K. B., & Sanders, S. (2004). Alzheimer’s caregiver differences in experience of loss, grief reactions and depressive symptoms across stage of disease: a mixed-method analysis. Dementia, 3(2), 195–210. Retrieved 2^nd^ November 2020, from: https://doi.org/10.1177/1471301204042337

Al-Janabi, H., Carmichael, F., & Oyebode, J. (2018). Informal care: choice or constraint? Scandinavian Journal ofCaring Sciences, 32(1), 157–167. Retrieved 27^th^ October 2020, from: https://doi.org/10.1111/scs.12441

Alzheimer’s Society. (2015). The progression of Alzheimer’s disease and other dementias. Retrieved 21^st^ January 2021, from: https://www.alzheimers.org.uk sites/default/files/pdf/factsheet_the_progression_of_alzheimers_disease_and_other dementias.pdf

Aminzadeh, F., Byszewski, A., Molnar, F. J., & Eisner, M. (2007). Emotional impact of dementia diagnosis: Exploring persons with dementia and caregivers’ perspectives. Aging & Mental Health, 11(3), 281–290. Retrieved 15^th^ March 2020, from: https://doi.org/10.1080/13607860600963695

Arcand, M. (2015). End-of-life issues in advanced dementia: Part 1: goals of care, decision-making process, andfamily education. Canadian Family Physician Medecin De Famille Canadien, 61(4), 330–334. Retrieved 10^th^ January 2021, from: https://pubmed.ncbi.nlm.nih.gov/25873700

Auer, S., & Reisberg, B. (1997). The GDS/FAST staging system. International Psychogeriatrics, 9(S1), 167–171. Retrieved 28^th^ February 2021, from: https://doi.org/10.1017/S1041610297004869

Bieber, A., Nguyen, N., Meyer, G., & Stephan, A. (2019). Influences on the access to and use of formal communitycare by people with dementia and their informal caregivers: a scoping review. BMC Health Services Research, 19(1), 88. Retrieved 27^th^ January 2021, from: https://doi.org/10.1186/s12913-018-3825-z

Blandin, K., & Pepin, R. (2017). Dementia grief: A theoretical model of a unique grief experience. Dementia(London), 16(1), 67–78. Retrieved 27^th^ January 2021, from: https://doi.org/10.1177/1471301215581081

Boss, P. (1999). Ambiguous loss: Learning to live with unresolved grief. Harvard University Press.

Brodaty, H., & Donkin, M. (2009). Family caregivers of people with dementia. Dialogues in Clinical Neuroscience,11(2), 217–228. Retrieved 20^th^ March 2020, from: https://pubmed.ncbi.nlm.nih.gov/19585957/

Brodaty, H., & Hadzi-Pavlovic, D. (1990). Psychosocial effects on carers of living with persons with dementia. Australian & New Zealand Journal of Psychiatry, 24(3), 351–361. Retrieved 20^th^ March 2020, from: https://doi.org/10.3109/00048679009077702

Brunton, M., Jordan, C., & Fouche, C. (2008). Managing public health care policy: who’s being forgotten? HealthPolicy, 88(2-3), 348–358. Retrieved 27^th^ January 2021, from: https://doi.org/10.1016/j.healthpol.2008.04.007

Chan, W. C., Ng, C., Mok, C.C. M., Wong, F. L. F., Pang, S. L., & Chiu, H. F. K. (2010). Lived experience of caregiversof persons with dementia in Hong Kong: a qualitative study. East Asian Archives of Psychiatry, 20(4), 163–168. Retrieved 12^th^ March 2020, from: https://search.informit.org/doi/10.3316/INFORMIT.934619447079403

Cheung, D.S.K., Ho, K.H.M., Cheung, T.F., Lam, S.C., Tse, M.M.Y. (2018). Anticipatory grief of spousal and adult children caregivers of people with dementia. BMC Palliat Care 17, 124. Retrieved 8^th^ January 2022, from: https://doi.org/10.1186/s12904-018-0376-3

Clemmensen, T. H., Busted, L. M., Søborg, J., & Bruun, P. (2016). The family’s experience and perception of phases and roles in the progression of dementia: An explorative, interview-based study. Dementia, 18(2),490–513. Retrieved 20^th^ March 2020, from: https://doi.org/10.1177/1471301216682602

Colling, K. B. (2004). Special section - behavioral symptoms of dementia: their measurement and intervention. Caregiver interventions for passive behaviors in dementia: links to the NDB model. Aging & MentalHealth, 8(2), 117–125. Retrieved 20^th^ March 2020, from: https://doi.org/10.1080/13607860410001649626

de la Cuesta-Benjumea, C. (2011). Strategies for the relief of burden in advanced dementia care-giving. Journal of Advanced Nursing, 67(8), 1790–1799. Retrieved 20^th^ March 2020, from: https://onlinelibrary.wiley.com/doi/abs/10.1111/j.1365-2648.2010.05607.x

Dementia Carers Count. (2019). Dementia care statistics: Key facts & figures. Retrieved 22nd February 2021, from:https://dementiacarers.org.uk/for-dementia-professionals/key-facts-figures/

De Silva, N., & Curzio, J. (2009). Transition into long-term care: the lived experience of carers of individuals withdementia. Mental Health Nursing, 29(5), 10–14. Retrieved 25^th^ March 2020, from: https://www.proquest.com/openview/63367e292b9e8c5f279df23c1dca883f/1?pq-origsite=gscholar&cbl=135348

Dupuis, S. L. (2002). Understanding ambiguous loss in the context of dementia care. Journal of GerontologicalSocial Work, 37(2), 93–115. Retrieved 18^th^ March 2020, from: https://doi.org/10.1300/J083v37n02_08

Evans, S., Waller, S., Bray, J., & Atkinson, T. (2019). Making homes more dementia-friendly through the use of aids and adaptations. Healthcare (Basel, Switzerland), 7(1), 43. Retrieved 17^th^ March 2020, from: https://doi.org/10.3390/healthcare7010043

Farina, N., Page, T. E., Daley, S., Brown, A., Bowling, A., Basset, T., Livingston, G., Knapp, M., Murray, J., & Banerjee, S. (2017). Factors associated with the quality of life of family carers of people with dementia: A systematic review. Alzheimer’s & Dementia,13(5), 572–581. Retrieved 17th March 2020, from: https://doi.org/https://doi.org/10.1016j.jalz.2016.12.010

Galvin, J. E., & Sadowsky, C. H. (2012). Practical guidelines for the recognition and diagnosis of dementia. The Journal of the American Board of Family Medicine, 25(3), 367. Retrieved 10^th^ January 2021, from: https://doi.org/10.3122%20jabfm.2012.03.100181

Givens, J. L., Prigerson, H. G., Kiely, D. K., Shaffer, M. L., & Mitchell, S. L. (2011). Grief among family members of nursing home residents with advanced dementia. American Journal of Geriatric Psychiatry, 19(6), 543–550. Retrieved 10^th^ January 2020, from: https://doi.org/10.1097/JGP.0b013e31820dcbe0

Greenwood, N., Pound, C., Smith, R., & Brearley, S. (2019). Experiences and support needs of older carers: A focusgroup study of perceptions from the voluntary and statutory sectors. Maturitas, 123, 40–44. Retrieved 10^th^ January 2021, from: https://doi.org/10.1016/j.maturitas.2019.02.003

Greenwood, N., & Smith, R. (2019). Motivations for being informal carers of people living with dementia: asystematic review of qualitative literature. BMC Geriatrics, 19(1), 169. Retrieved 10^th^ January 2021, from: https://doi.org/10.1186%20s12877-019-1185-0

Hall, M., & Sikes, P. (2018). From “What the Hell Is Going on?” to the “Mushy Middle Ground” to “Getting Used toa New Normal”: Young people’s biographical narratives around navigating parental dementia. Illness, Crises & Loss, 26(2), 124–144. Retrieved 10^th^ October 2020, from: https://doi.org/10.1177/1054137316651384

Harris, D. (2007). Forget me not: palliative care for people with dementia. Postgraduate Medical Journal, 83(980), 362–366. Retrieved 10th January 2021, from: https://doi.org/10.1136/pgmj.2006.052936

Hawker, S., Payne, S., Kerr, C., Hardey, M., & Powell, J. (2002). Appraising the evidence: Reviewing disparate datasystematically. Qualitative Health Research, 12(9), 1284–1299. Retrieved 10th October 2020, from: https://doi.org/10.1177%201049732302238251#

Hellström, I., Nolan, M., & Lundh, U. (2005). ‘We do things together’: A case study of ‘couplehood’ in dementia. Dementia, 4(1), 7–22. Retrieved 10th January 2020, from: https://doi.org/10.1177/1471301205049188

Idura, A. M., Noorlaili, M. T., Rosdinom, R., Azlin, B., & Iryani, M. D. (2018). Caring for moderate to severe dementia patients - Malaysian family caregivers experience. International Medical Journal Malaysia,17(1), 101–110.

James, C.O. (2021). Moral Distress in the Care of People Living with Moderate to Advanced Dementia: A Narrative Exploration of Family Carers’ Experience of Home-Based Care Provision towards the End of Life. PhD Thesis, Lancaster University.

James, C., Walshe, C., & Froggatt, K. (2020). Protocol for a systematic review on the experience of informal caregivers for people with a moderate to advanced dementia within a domestic home setting. Systematic Reviews, 9(1), 270. Retrieved 28^th^ February 2021, from: https://doi.org/10.1186/s13643-020-01525-0

Jameton, A. (1984). Nursing practice: The ethical issues. Prentice-Hall.

Jameton, A. (1993). Dilemmas of moral distress: moral responsibility and nursing practice. AWHONNS Clinical Issues in Perinatal and Women’s Health Nursing, 4(4), 542–551. Retrieved 28^th^ February 2019, from: https://pubmed.ncbi.nlm.nih.gov/8220368/

Joling, K. J., Janssen, O., Francke, A. L., Verheij, R. A., Lissenberg-Witte, B. I., Visser, P. J., & van Hout, H. P. (2020). Time from diagnosis to institutionalization and death in people with dementia. Alzheimer’s & Dementia,16(4), 662–671. Retrieved 28^th^ January 2021, from: https://doi.org/10.1002/alz.12063

Jones, P. S., & Martinson, I. M. (1992). The experience of bereavement in caregivers of family members with Alzheimer’s disease. Image Journal of Nursing Scholarship, 24(3), 172–176. Retrieved 28th January 2021, from: https://doi.org/10.1111j.1547-5069.1992.tb00714.x

Karg, N., Graessel, E., Randzio, O., & Pendergrass, A. (2018). Dementia as a predictor of care-related quality of lifein informal caregivers: a cross-sectional study to investigate differences in health-related outcomes between dementia and non-dementia caregivers. BMC Geriatrics, 18(1), 189. Retrieved 28th January 2021, from: https://doi.org/10.1186/s12877-018-0885-1

Karger, C. R. (2018). Emotional experience in patients with advanced Alzheimer’s disease from the perspective offamilies, professional caregivers, physicians,and scientists. Aging & Mental Health, 22(3), 316–322.

Knapp, M., Prince, M., Guerchet, M., McCrone, P., Prina, M., Comas-Herrera, A., Wittenberg, R., Adelaja, B., Hu, B.,King, D., Rehill, A., & Salimkumar, D. (2014). Dementia UK: Update (second edition).

Kumar, A. S., J.; Goyal, A.; Tsao, J.W. (2020). Alzheimer disease. Stat Pearls Publishing; 2020. Retrieved 8th January 2021, from: https://www.ncbi.nlm.nih.gov/books/NBK499922/

Kwon, Y., Lemieux, M., McTavish, J., & Wathen, N. (2015). Identifying and removing duplicate records from systematic review searches. Journal of the Medical Library Association: JMLA, 103(4), 184–188. Retrieved 15^th^ January 2021, from: https://doi.org/10.3163/1536-5050.103.4.004

Lee, L., & Weston, W. W. (2011). Disclosing a diagnosis of dementia: helping learners to break bad news. Canadian Family Physician Medecin De Famille Canadien, 57(7), 851–e272. Retrieved 25^th^ January 2020, from: https://pubmed.ncbi.nlm.nih.gov/21753111

Lethin, C., Hallberg, I. R., Karlsson, S., & Janlöv, A. (2016). Family caregivers experiences of formal care when caring for persons with dementia through the process of the disease. Scandinavian Journal of Caring Sciences, 30(3), 526–534. Retrieved 27^th^ March 2021, from: https://doi.org/10.1111/scs.12275

Lewis, F., Sussex, J., Schaffer, S. K., O’Neill, P., & Cockcroft, L. (2014). The trajectory of dementia in the UK - Making a difference. IDEAS Working Paper Series from RePEc. Retrieved 15^th^ February 2020, from: https://www.ohe.org/system/files/private/publications/401%20-%20Trajectory_dementia_UK_2014.pdf

Lewis, L. F. (2014). Caregivers’ experiences seeking hospice care for loved ones with dementia. Qualitative Health Research, 24(9), 1221–1231. Retrieved 15th February 2020, from: https://doi.org/10.1177/1049732314545888

Lindauer, A., Harvath, T. A., Berry, P. H., & Wros, P. (2016). The meanings African American caregivers ascribe todementia-related changes: The paradox of hanging on to loss. Gerontologist, 56(4), 733–742. Retrieved 15^th^ February 2020, from: https://www.ncbi.nlm.nih.gov/pmc/articles/PMC6282689/pdf/gnv023.pdf

Lindeza, P., Rodrigues, M., Costa, J., Guerreiro, M., & Rosa, M. M. (2020). Impact of dementia on informal care: asystematic review of family caregivers’ perceptions. BMJ Supportive & Palliative Care, bmjspcare-2020-002242. Retrieved 15^th^ February 2021, from: https://doi.org/10.1136/bmjspcare-2020-002242

Lorenc, T., Petticrew, M., Whitehead, M., Neary, D., Clayton, S., Wright, K., Thomson, H., Cummins, S., Sowden, A., & Renton, A. (2014). Appendix 5: Quality assessment for the systematic review of qualitative evidence. InCrime, fear of crime and mental health: synthesis of theory and systematic reviews of interventions and qualitative evidence. NIHR Journals Library. Retrieved 25^th^ January 2020, from: https://doi.org/10.3310/phr02020

Lorenzetti, D. L., & Ghali, W. A. (2013). Reference management software for systematic reviews and meta-analyses: an exploration of usage and usability. BMC Med Res Methodol, 13, 141. Retrieved 25^th^ January 2020, from: https://doi.org/10.1186/1471-2288-13-141

Marshall, G. A., Amariglio, R. E., Sperling, R. A., & Rentz, D. M. (2012). Activities of daily living: where do they fit inthe diagnosis of Alzheimer’s disease? Neurodegenerative Disease Management, 2(5), 483–491. Retrieved 25^th^ August 2020, from: https://doi.org/10.2217/nmt.12.55

Mesterton, J., Wimo, A., By, A., Langworth, S., Winblad, B., & Jönsson, L. (2010). Cross sectional observationalstudy on the societal costs of Alzheimer’s disease. Current Alzheimer Research, 7(4), 358–367. Retrieved 23^rd^ January 2021, from: https://doi.org/10.2174/156720510791162430

Meuser, T. M., & Marwit, S. J. (2001). A comprehensive, stage-sensitive model of grief in dementia caregiving. TheGerontologist, 41(5), 658–670. Retrieved 13^th^ June 2020, from: https://doi.org/10.1093/geront/41.5.658

Mlinac, M. E., & Feng, M. C. (2016). Assessment of activities of daily living, self-care, and independence. Archives of Clinical Neuropsychology, 31(6), 506–516. Retrieved 17^th^ January 2021, from: https://doi.org/10.1093/arclin/acw049

Moore, K. J., Davis, S., Gola, A., Harrington, J., Kupeli, N., Vickerstaff, V., King, M., Leavey, G., Nazareth, I., Jones, L.,& Sampson, E. L. (2017). Experiences of end of life amongst family carers of people with advanced dementia: longitudinal cohort study with mixed methods. BMC Geriatrics, 17(135).

Oh, Y., & Gastmans, C. (2015). Moral distress experienced by nurses: A quantitative literature review. 22(1), 15–31. Retrieved 18^th^ October 2020, from: https://doi.org/10.1177/0969733013502803

Page, M. J., McKenzie, J.E., Bossuyt, P.M., Boutron, I., Hoffmann, T.C., Mulrow, C.D., et al. (2020). The PRISMA 2020 statement: an updated guideline for reporting systematic reviews. BMJ 2021; 372. Retrieved 15^th^ July 2021, from: https://doi.org/10.1136/bmj.n160

Pérez-González, A., Vilajoana-Celaya, J, Guàrdia-Olmos, J. (2021). Alzheimer’s Disease Caregiver Characteristics and Their Relationship with Anticipatory Grief. International Journal of Environmental Research and Public Health. 18(16):8838. Retrieved 8^th^ January 2022, from: https://doi.org/10.3390/ijerph18168838

Pertl, M. M., Sooknarine-Rajpatty, A., Brennan, S., Robertson, I. H., & Lawlor, B. A. (2019). Caregiver choice and caregiver outcomes: A longitudinal study of Irish spousal dementia caregivers. Frontiers in Psychology, 10(1801). Retrieved 18^th^ October 2020, from: https://doi.org/10.3389/fpsyg.2019.01801

Phillips, J., Pond, D., & Goode, S. M. (2011). Timely diagnosis of dementia: Can we do better? A report for Alzheimer’s Australia. Alzheimer’s Australia. Paper 24. Retrieved 18^th^ October 2020, from: https://www.dementia.org.au/sites/default/files/Timely_Diagnosis_Can_we_do_better.pdf

Polit, D. F., & Beck, C. T. (2010). Generalization in quantitative and qualitative research: Myths and strategies.International Journal of Nursing Studies, 47(11), 1451–1458. Retrieved 18^th^ October 2020, from: https://doi.org/10.1016%20j.ijnurstu.2010.06.004

Popay, J., Roberts, H., Sowden, A., Petticrew, M., Arai, L., Rodgers, M., Britten, N., Roen, K., & Duffy, S. (2006). Guidance on the conduct of narrative synthesis in systematic Reviews. Retrieved 18^th^ March 2020, from: https://www.lancaster.ac.uk/media/lancaster-university/content-assets/documents/fhm/dhr/chir/NSsynthesisguidanceVersion1-April2006.pdf

Prigerson, H. G., & Maciejewski, P. K. (2008). Grief and acceptance as opposite sides of the same coin: setting a research agenda to study peaceful acceptance of loss. British Journal of Psychiatry, 193(6), 435–437. Retrieved 18^th^ March 2020, from: https://doi.org/10.1192/bjp.bp.108.053157

Rando, T. A. (2000). Clinical dimensions of anticipatory mourning: Theory and practice in working with the dying,their loved ones, and their caregivers. Research Press.

Reinhard, S., Levine, C., & Samis, S. (2012). Home alone: Family caregivers providing complex chronic care. Retrieved 18^th^ January 2021, from: https://www.aarp.org/content/dam/aarp/research/public_policy_institute/health/home-alone-family-caregivers-providing-complex-chronic-care-rev-AARP-ppi-health.pdf

Sanders, A. E. (2016). Caregiver stress and the patient with dementia. Continuum: Lifelong Learning in Neurology, 22(2), 619–625. Retrieved 27^th^ March 2020, from: https://doi.org/10.1212/con.0000000000000301

Schmidt, K. (2014). Clinical dementia rating scale. In A. C. Michalos (Ed.), Encyclopedia of Quality of Life andWell-Being Research (pp. 957–960). Springer Netherlands. Retrieved 20^th^ January 2021, from: https://doi.org/10.1007978-94-007-0753-5_690

Schulz, R., Beach, S. R., Cook, T. B., Martire, L. M., Tomlinson, J. M., & Monin, J. K. (2012). Predictors and consequences of perceived lack of choice in becoming an informal caregiver. Aging & Mental Health,16(6), 712–721. Retrieved 27^th^ January 2021, from: https://doi.org/10.1080/13607863.2011.651439

Schulz, R., & Eden, J. (2016). Family caregiving roles and impacts. In R. Schulz & J. Eden (Eds.), Families Caring for an Aging America. National Academies Press (US). Retrieved 27^th^ January 2021, from: https://doi.org/10.17226/23606

Shuter, P., Beattie, E., & Edwards, H. (2014). An exploratory study of grief and health-related quality of life forcaregivers of people with dementia. 29(4), 379–385. Retrieved 21^st^ July 2020, from: https://doi.org/10.1177/1533317513517034

Silén, M. (2011). Encountering ethical problems and moral distress as a nurse (Publication Number 20), JönköpingUniversity.

Smith, A. L., Lauret, R., Peery, A., & Mueller, T. (2001). Caregiver needs: a qualitative exploration. Clinical Gerontologist, 24(1), 3–26. Retrieved 20^th^ March 2020, from: https://doi.org/10.1300/J018v24n01_02

Swall, A, Williams, C, Marmstål Hammar, L. (2020). The value of “us”—Expressions of togetherness in couples where one spouse has dementia. International Journal of Older People Nursing. 15:e12299. Retrieved 4^th^ April 2021, from: https://doi.org/10.1111/opn.12299

Tretteteig, S., Vatne, S., & Rokstad, A. M. (2017). The influence of day care centres designed for people withdementia on family caregivers - a qualitative study. BMC Geriatrics, 17(1), 5. Retrieved 20^th^ March 2020, from: http://dx.doi.org/10.1186/s12877-016-0403-2

Tretteteig, S., Vatne, S., & Rokstad, A. M. (2017). Meaning in family caregiving for people with dementia: a narrative study about relationships, values, and motivation, and how day care influences these factors.Journal of Multidisciplinary Healthcare, 10, 445–455. Retrieved 20^th^ March 2020, from: https://doi.org/10.2147/JMDH.S151507

van Gennip, I.E., Roeline, H., Pasman, W., Oosterveld-Vlug, M.G., Willems, D.L., Onwuteaka-Philipsen, B.D. (2016). How Dementia Affects Personal Dignity: A Qualitative Study on the Perspective of Individuals with Mild to Moderate Dementia. The Journals of Gerontology: Series B, 71(3), 491–501, Retrieved 4^th^ January 2022, from: https://doi.org/10.1093/geronb/gbu137

van Wijngaarden, E., van der Wedden, H., Henning, Z., Komen, R., & The, A. (2018). Entangled in uncertainty: Theexperience of living with dementia from the perspective of family caregivers. PLOS ONE, 13(6), e0198034–e0198034. Retrieved 12^th^ February 2021, from: https://doi.org/10.1371/journal.pone.0198034

Vestergaard, A.H.S., Sampson, E.L., Johnsen, S.P., Petersen, I. (2020). Social Inequalities in Life Expectancy and Mortality in People with Dementia in the United Kingdom. Alzheimer Disease & Associated Disorders. 34(3), 254–261. Retrieved 4^th^ January 2022, from: https://doi.org/10.1097/WAD.0000000000000378

Wladkowski, S. P. (2016). Live discharge from hospice and the grief experience of dementia caregivers. Journal of Social Work. In: End-of-Life & Palliative Care, 12(1), 47–62. Retrieved 5th March 2020, from: https://doi.org/10.1080/15524256.2016.1156600

Woodman, C., Baillie, J., & Sivell, S. (2016). The preferences and perspectives of family caregivers towards place ofcare for their relatives at the end-of-life. A systematic review and thematic synthesis of the qualitative evidence. BMJ Supportive & Palliative Care, 6(4), 418. Retrieved 27^th^ May 2020, from: https://doi.org/10.1136bmjspcare-2014-000794

Xanthopoulou, P., McCabe, R. (2019). Subjective experiences of cognitive decline and receiving a diagnosis of dementia: qualitative interviews with people recently diagnosed in memory clinics in the UK. BMJ Open 2019;9: e026071. Retrieved 4^th^ January 2022, from: https://doi.org/10.1136/bmjopen-2018-026071

