## Supplementary Table 1: Result of the quality assessment of included studies for "The experience of family carers for people with moderate to advanced dementia within a domestic home setting: a systematically constructed narrative synthesis"

| Author | Abstract/title | Introduction /aims | Data collection | Sampling | Analysis | Ethics/ Bias | Results/ Findings | Generalisability | Implication | Total | Grade |
|---|---|---|---|---|---|---|---|---|---|---|---|
| **Adams & Sanders (2004)** | 3 | 4 | 4 | 4 | 4 | 2 | 3 | 2 | 3 | 29 | B |
| **Brunton et al. (2008)** | 3 | 4 | 4 | 4 | 4 | 4 | 4 | 2 | 4 | 34 | A |
| **Chan et al. (2010)** | 4 | 4 | 4 | 4 | 4 | 4 | 4 | 4 | 4 | 36 | A |
| **Colling (2004)** | 3 | 4 | 4 | 4 | 4 | 4 | 4 | 4 | 4 | 35 | A |
| **de La Cuesta-Benjumea (2011)** | 4 | 4 | 4 | 4 | 4 | 4 | 4 | 4 | 4 | 36 | A |
| **De Silva and Curzio (2009)** | 4 | 4 | 3 | 1 | 3 | 4 | 4 | 2 | 4 | 29 | B |
| **Hirschman et al. (2006)** | 3 | 4 | 4 | 4 | 4 | 4 | 4 | 4 | 4 | 35 | A |
| **Idura et al. (2018)** | 4 | 3 | 4 | 4 | 4 | 4 | 4 | 4 | 4 | 35 | A |
| **Karger (2018)** | 4 | 4 | 4 | 4 | 4 | 1 | 4 | 4 | 4 | 32 | A |
| **Lamahewa et al. (2018)** | 4 | 4 | 4 | 2 | 4 | 4 | 4 | 2 | 4 | 32 | A |
| **Lethin et al. (2016)** | 4 | 4 | 4 | 4 | 4 | 4 | 4 | 4 | 4 | 36 | A |
| **Lindauer et al. (2016)** | 4 | 4 | 4 | 4 | 4 | 4 | 4 | 4 | 4 | 36 | A |
| **Moore et al. (2017)** | 4 | 4 | 4 | 4 | 4 | 4 | 4 | 4 | 4 | 36 | A |
| **Robinson et al. (2010)** | 3 | 4 | 4 | 4 | 4 | 4 | 4 | 4 | 4 | 35 | A |
| **Smith et al. (2001)** | 3 | 4 | 4 | 4 | 4 | 4 | 4 | 4 | 4 | 35 | A |
| **Tretteteig et al. (2017)** | 4 | 4 | 4 | 3 | 4 | 4 | 4 | 4 | 4 | 35 | A |
| **Wladkowski (2016)** | 3 | 4 | 4 | 4 | 4 | 4 | 4 | 4 | 4 | 35 | A |
