## Supplementary Table 2: Textual narrative synthesis exploring relationships between studies for "The experience of family carers for people with moderate to advanced dementia within a domestic home setting: a systematically constructed narrative synthesis"

| **Stage** | **Country** | **Authors and combined population across all locations and participants type** (Family caregivers: including spouses, adult children, in-laws and other relatives) | **Common focus between studies** |
|---|---|---|---|
| **Moderate to advanced stage** | USA | Adams and Sanders (2004) *[moderate (n=41), advanced (n=33)]* | Determinants of how the experience of loss, burden and grief reactions symptoms are expressed at the moderate stage and how caregivers make sense of dementia-related changes |
| | | Colling (2004) *[moderate (n=16), advanced (n=19)]* | |
| | | Smith et al. (2001) *[moderate (n=21), advanced (n=11)]* | |
| | | Lindauer et al. (2016) *[Total participants (n= 11), specific stage breakdown was not given in paper]* | |
| | Hong Kong | Chan et al. (2010); *[Total participants (n= 27), stage breakdown was not given in paper]* | |
| | Norway | Tretteteig et al. (2017) *[moderate (n=2), advanced (n=2)]* | Determinants of factors that influence the caregivers' perception of their own needs and motivation in the role of caring for their family members |
| | Malaysia | Idura et al. (2018) *[moderate (n=6), advanced (n=6)]* | |
| **Advanced stage** | Hong Kong | Chan et al., (2010) *[Total participants (n= 27), specific stage breakdown was not given in paper]* | Determinants of how the experience of loss, burden and grief reactions symptoms are expressed at the advanced or severe stage and how caregivers make medical decisions and make |
| | USA | Adams and Sanders, (2004) *[advanced (n=33)]* | |

| Stage | Country | **Authors and combined population across all locations and participants type** (Family caregivers: including spouses, adult children, in-laws and other relatives) | **Common focus between studies** |
|---|---|---|---|
| | | Colling (2004) *[advanced (n=19)]*<br><br>Smith et al., (2011) *[advanced (n=11)]*<br><br>Hirschman et al. (2006) *[advanced (n=30)]*<br><br>Lindauer et al. (2016) *[Total participants (n= 11), specific stage breakdown was not given in paper]* | sense of dementia-related changes |
| | UK | De Silva and Curzio, (2010) *[advanced (n=10)]*<br><br>Lamahewa et al., (2018) *[advanced (n=10)]*<br><br>Moore et al., (2017) *[advanced (n=6)]*<br><br>Wladkowski (2016) *[advanced (n=24)]* | |
| | Germany | Karger (2018) *[advanced (n= 20)]* | Factors that influence caregivers' emotional response to the grieving process/impact on caregivers' grieving process and influence decision-making strategies at the advanced stage/end of life |
| | Sweden | Lethin et al. (2016) *[advanced*<br><br>*(n= 23)]* | Factors that influence how caregivers' view formal care for their relatives during advanced dementia |
| | Canada | Robinson et al., (2010) *[advanced (n= 29)]* | |

| **Stage** | **Country** | **Authors and combined population across all locations and participants type** (Family caregivers: including spouses, adult children, in-laws and other relatives) | **Common focus between studies** |
|---|---|---|---|
| | New Zealand | Brunton et al. (2008) *[Total participants* <br><br> *(n= 5), specific stage breakdown was not given in paper]* | Determinants of the need for rest and coping strategies in caregivers' role at the advanced stage. |
| | Spain | de La Cuesta-Benjumea, (2011) *[advanced (n= 23)]* | |
