## Supplementary Table 3: Preliminary synthesis of studies for "The experience of family carers for people with moderate to advanced dementia within a domestic home setting: a systematically constructed narrative synthesis"

| | |
|---|---|
| **Design** | |
| Cross-sectional | Adams and Sanders (2004) |
| Longitudinal cohort | Moore et al. (2017) |
| Focus group | Chan et al. (2010); Karger (2018); Lamahewa et al. (2018); Lethin et al. (2016); Robinson et al. (2010) |
| Phenomenology (including Interpretative Phenomenological Approach) | Colling (2004); De Silva and Curzio (2009); Lindauer et al. (2016) |
| Qualitative | Hirschman et al. (2006); Idura et al. (2018); Wladkowski (2016) |
| Grounded theory | de La Cuesta-Benjumea (2011) |
| Exploratory | Brunton et al. (2008) |
| Ethnography | Smith et al. (2011) |
| Narrative (Case study) | Tretteteig et al. (2017) |
| **Population** | |
| Family caregivers (generic category including spouses, adult children, in-laws and other relatives) | Adams and Sanders (2004); Brunton et al. (2008); Chan et al. (2010); Colling (2004); De Silva and Curzio (2009); Hirschman et al. (2006); Idura et al. (2018); Karger (2018); Lamahewa et al. (2018); Lethin et al. (2016); Lindauer et al. (2016); Moore et al. (2017); Robinson et al. (2010); Tretteteig et al. (2017); Wladkowski (2016) |
| Family caregiver (spouses only) | de La Cuesta-Benjumea (2011) |
| **Dementia stage** | |
| Advanced or severe | Brunton et al. (2008); de La Cuesta-Benjumea (2011); De Silva and Curzio (2009); Hirschman et al. (2006); Karger (2018); Lamahewa et al. (2018); Lethin et al. (2016); Moore et al. (2017); Robinson et al. (2010); Smith et al. (2001); Wladkowski (2016) |
| Moderate to advanced or severe (Combined accounts) | Adams and Sanders (2004); Chan et al. (2010); Colling (2004); Idura et al. (2018); Lindauer et al. (2016); Tretteteig et al. (2017) |
| **Location** | |
| United Kingdom (UK) | De Silva and Curzio (2009); Lamahewa et al. (2018); Moore et al. (2017); Wladkowski (2016) |
| United States of America (USA) | Adams and Sanders (2004); Colling (2004); Hirschman et al. (2006); Lindauer et al. (2016); Smith et al. (2001) |
| Norway | Tretteteig et al. (2017) |
| Sweden | Lethin et al. (2016) |
| Hong Kong | Chan et al. (2010) |
| New Zealand | Brunton et al. (2008) |
| Spain | de La Cuesta-Benjumea (2011) |
| Germany | Karger (2018) |
| Canada | Robinson et al. (2010) |
| Malaysia | Idura et al. (2018) |
